# Public involvement and co-design of longitudinal studies of sleep health alongside young people with rare genetic conditions

**DOI:** 10.64898/2026.04.07.26348880

**Authors:** Julie P. Clayton, Josephine E Haddon, Jessica Hall, Meg Attwood, Christopher Jarrold, Lioba C. S. Berndt, Abiola Saka, Marianne B.M. van den Bree, Matt W. Jones, Collaboration: Sleep Detectives Lived Experience Advisory Panel

## Abstract

**Background:** The mechanisms underpinning associations between sleep and psychiatric conditions are poorly understood, partly due to challenges with longitudinal sleep studies outside the laboratory. Children and young people with rare genetic conditions caused by micro-deletions or -duplications (Copy Number Variants or CNVs) have increased risk of disrupted sleep and poorer neurodevelopmental (ND) outcomes. The ‘Sleep Detectives’ study aims to investigate this by tracking behavioural and neurophysiological signatures of sleep health in young people with ND risk or ND-CNVs. To optimally achieve this, we have worked with families with ND-CNVs and charity partners to co-design our tools, methods, study protocol, and materials.

**Method:** We established a Lived Experience Advisory Group (LEAP) with nine parents and 13 children and young people with ND-CNVs, alongside representatives of UK charities Max Appeal and Unique. Together, the research team and LEAP co-designed two in-person family workshops in which we collected feedback on the acceptability of sleep monitoring devices, the design of bespoke cognitive tasks, and overall study protocol. Informal interviews and surveys were conducted with LEAP members and researchers, to enable the team to reflect and learn from their Patient/Public Involvement (PPI) experiences.

**Results:** Key outputs included pre-workshop invitation and briefing materials and insights that iteratively refined the main study design, including the need for flexibility to increase accessibility, selection of sleep devices, customisation of cognitive tasks, and choice of language in documents. The PPI process was highly valued by LEAP members, workshop attendees, and the research team. One investigator described the PPI work as “reinvigorating my love of research by helping me focus on science that matters”. Participating families also established peer support networks.

**Conclusions:** Involving families affected by ND-CNVs in co-designing the Sleep Detectives study maximised opportunities for acceptability, accessibility and scalability. The research team gained inspiration and deeper understanding of the impact of ND-CNVs on families. Families gained awareness about research, established connections with each other and peer support, and were enthusiastic about future research involvement. This experience empowered families to engage more deeply with the research process and helped the PPI work to be more impactful and inclusive.

**Plain English summary:** Children and young people with rare genetic conditions caused by small deletion or duplication of genetic material are more likely to experience sleep difficulties such as insomnia, restless sleep, and tiredness. They also show an increased likelihood of neurodevelopmental conditions such as learning disability and autism, and mental health issues such as anxiety. The Sleep Detectives team wanted to explore how these genetic conditions affect children’s sleep, cognition and psychiatric health. To make sure that the project design was well suited to the children and young people that would be invited to participate, the team worked closely with families to design the study. Parents and caregivers of affected children and young people were invited to join a Lived Experience Advisory Panel (LEAP), together with charity representatives and Sleep Detective researchers, to co-design two hands-on workshops, and advise on study design. Children and young people and parents/caregivers attending the workshops tried out and provided feedback on tools and devices that the research team were developing. They also advised on the arrangements and support families might need whilst taking part, and on the study protocol. This collaborative approach helped ensure the study design was optimally suited for the recruitment and participation of children and young people and their families. This report documents our public involvement work for the Sleep Detectives study, illustrating the difference the partnership between researchers and families has made to the project, and the wider benefits for all concerned.

## Background

Sleep Detectives is a Wellcome Trust-funded study in the UK which seeks to understand and explain the relationships between sleep difficulties, cognition, and psychiatric health in children and young people with rare genetic conditions caused by micro-deletions or - duplications in DNA (Copy Number Variants, CNVs). Our focus is on CNVs associated with a high likelihood of neurodevelopmental conditions (ND-CNVs) [1,2] including 22q11.2 deletions, 1q21.1 deletions and 16p11.2 duplications. Those affected by these CNVs are more likely to experience cognitive impairment [3], mental health conditions [4] and poor sleep [5,6].

Throughout the study, guided by the principle of ‘nothing about us, without us’ - which originated in the disability rights movement [7] – and by Wellcome policy, the Sleep Detectives study has sought to create a research environment and participatory processes that support and encourage children and young people with ND-CNVs and their families.

This approach has centred on enabling them to share their perspectives, have their voices heard, and contribute meaningfully to the co-design of the study. It sought to elicit their views on the practical and accessible aspects of study design, including what felt feasible in terms of participation, communication formats, and support needs. This way, we hoped to empower public contributors to co-design the research study: to share decision-making, improve research design and implementation [8], as well as promote transparency around the study’s relevance and our ethical accountability. This approach aimed to ensure that lived experience informed the development of our research priorities, methods, and materials throughout. Sleep Detectives goes beyond previous studies in aiming to measure sleep over prolonged periods, initially conducting measurements over 8 days and nights, as a step towards multi-year longitudinal studies. Given the possible additional challenges that repeat monitoring may have for participating families, the involvement of families in designing the study was even more important for ensuring feasibility and acceptability.

### The importance of healthy sleep

Sleep is crucial for nearly all aspects of physical and mental health in a way that varies across the life course. In adults, sleep helps consolidate memories [9,10] and process emotions [11], supports immune function [12,13] and cardiometabolic health, and is tightly linked with our mental health and functioning [14]. There are bidirectional interrelationships between sleep and psychiatric symptoms, whereby disrupted or poor-quality sleep is associated with increases in perceptual differences, anxiety, irritability and, in more rare instances, other unusual experiences such as a sense of feeling detached from oneself; seeing, hearing, or sensing things that others around you may not experience; or strongly held beliefs that may not align with how others interpret the situation. Conversely, periods of better sleep generally improve these symptoms [15]. During childhood and adolescence, sleep also affects neurodevelopment and the associated emergence and refinement of neural circuits [16,17]. As such, early-life sleep disruption can have lasting consequences for lifelong mental health [18,19].

### Sleep in rare genetic conditions caused by ND-CNVs

Sleep is commonly disrupted in children and young people with ND-CNVs, who experience high rates of fatigue and sleep fragmentation, with 41% showing symptoms of insomnia from an average age of 3 years [20,21]. These sleep problems have been linked to behavioural, emotional and psychological problems [5,21,22,23], and may indicate those who are at greater risk of psychiatric, cognitive and motor coordination difficulties [5]. Detailed electroencephalogram (EEG) studies of sleep architecture show that children and young people with a CNV on chromosome 22, causing 22q11.2 deletion syndrome (22q11.2DS), exhibit objective differences compared to unaffected siblings. Specifically, they show an increased proportion of N3 (slow-wave) sleep, alongside atypical patterns in sleep spindle density and slow-wave characteristics. These oscillatory features—spindles and slow waves—reflect circuit mechanisms of memory consolidation and are also atypical in adults diagnosed with schizophrenia [24] or depression [25]. Whilst in neurotypical siblings of children and young people with ND-CNV, higher spindle and slow-wave amplitudes correlate positively with overnight memory performance, in individuals with 22q11.2DS, a negative correlation has been observed, suggesting disrupted interrelationships between sleep physiology and cognitive function in 22q11.2DS [21].

### The Sleep Detectives

The features of sleep detected by EEG, notably the properties and coordination of spindle and slow wave oscillations, are promising measurable indicators (biomarkers) of brain dysfunction in neuropsychiatric conditions [26,27,28]. However, the relationship between genetic risk, sleep difficulties, and the development of neuropsychiatric problems remains to be fully characterised, particularly over timescales spanning childhood and adolescence. The Sleep Detectives study seeks to close this gap by examining how behavioural and neurophysiological signatures of sleep health are associated with impaired cognition and neuropsychiatric risk in children and young people with ND-CNV and their unaffected siblings (without ND-CNV). The Sleep Detectives study aims to optimise a pipeline to study sleep in children and young people with ND-CNVs using low-burden tools and repeatable, at-home methods of measuring sleep and sleep-sensitive cognition. This will be achieved using three types of assessment, from nearable devices (such as bedside monitors), from wearable devices (like wrist-worn actigraphy and low-density EEG), and from custom-designed cognitive tasks.

This paper reports on our Patient/Public Involvement (PPI) and co-design process and its influence on the Sleep Detectives study design, implementation, and outcomes. Our PPI aims were to:

- establish a Lived Experience Advisory Panel (LEAP);
- work with the LEAP to co-design and deliver two engaging, hands-on workshops with families with children or young people with ND-CNVs in which we received feedback on the acceptability of potential sleep monitoring devices, bespoke cognitive tasks, and feasibility of the overall study protocol;
- incorporate, where possible, feedback from the workshops and LEAP meetings into the overall study design and maintain communications during data acquisition.

## Methods

### Study

The PPI work reported here forms part of a research project (Wellcome Grant 226709/Z/22/Z) to investigate sleep in young people with rare genetic conditions associated with high likelihood of psychiatric disorders. The project is led by researchers from University of Bristol, Cardiff University, and University of Exeter, with a dedicated co-investigator with expertise in PPI (JC), leading on strategic planning, coordination and delivery of PPI activities. This includes liaising with charities to identify and recruit families with lived experience of ND-CNVs and setting up and managing PPI activities, ensuring that they are accessible, meaningful, and well-supported.

### Ethical approval

Ethical approval for the Sleep Detectives project has been received from the University of Bristol Faculties of Life Sciences and Science Research Ethics Committee (17737, for a pilot with neurotypical children recruited through school) and the Cardiff University School of Medicine Research Ethics Committee (SREC reference: 25/14), for the main study in children and young people with a ND-CNV and their unaffected siblings (without a ND-CNV). The Lived Experience Advisory Panel (LEAP) and PPI workshops brought together charity partners, parents, and children and young people with ND-CNV. Formal written consent was not sought for PPI participation, as workshop attendees were engaged in an advisory capacity only, rather than as research participants. Ethics approval was not applicable for these PPI advisory-only activities. This is in accordance with best practice standards of the UK National Institute for Health and Care Research which state that ethics approval is not required for PPI work https://www.nihr.ac.uk/reporting-ppi-publications-guidance. Verbal consent was given prior to the workshops commencing, and comprehensive information about the research process was provided by the PPI lead and researchers to ensure transparency and informed involvement.

### Setting up PPI and co-design processes

The three stages of the Sleep Detectives PPI work are summarised in **Figure 1**.

**Figure 1.**
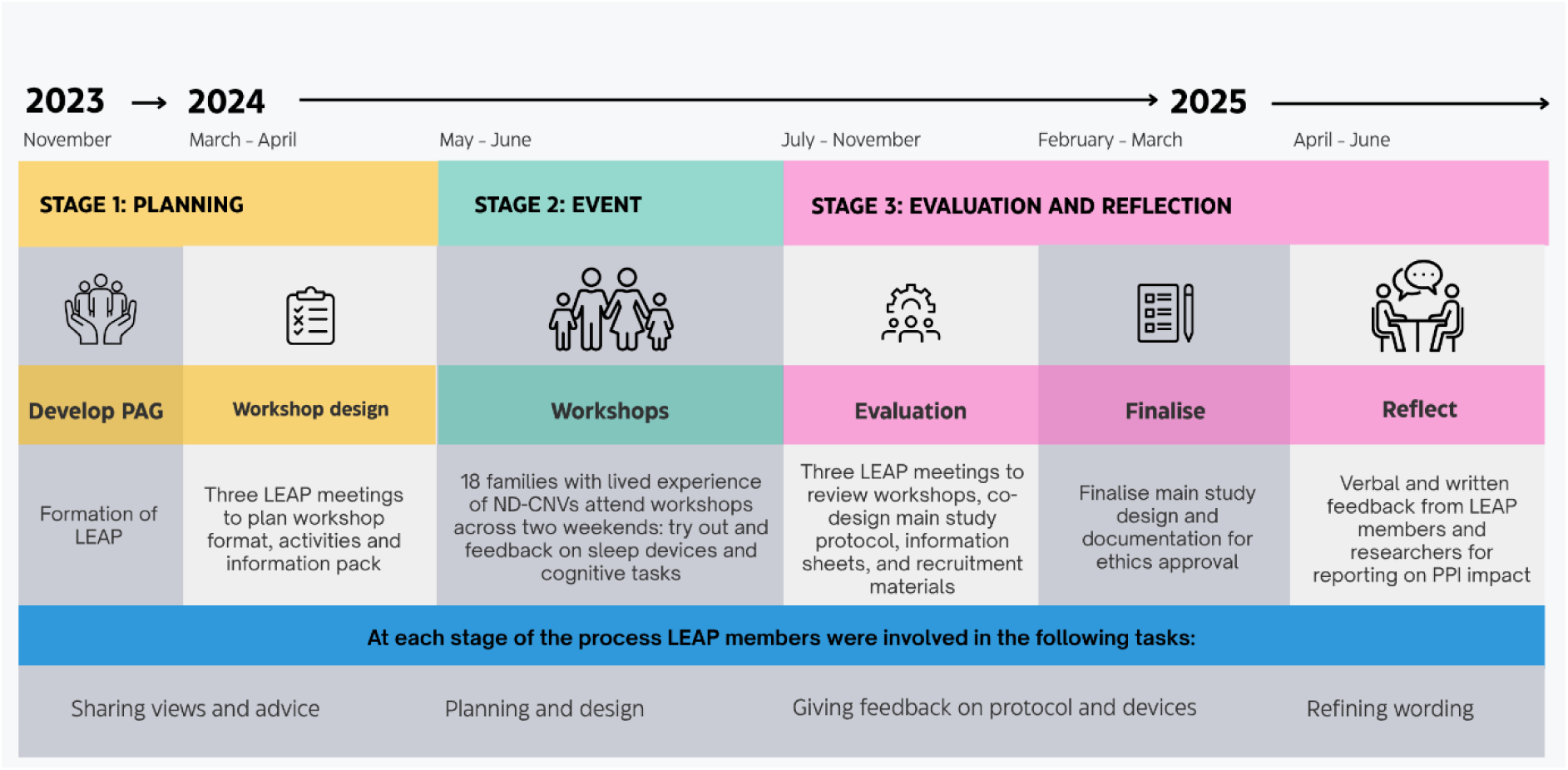
Sleep Detectives public involvement timeline – key dates and activities Legend: Patient/Public Involvement (PPI) in Sleep Detectives was conducted through three stages, Planning, Event and Evaluation, and Reflection, and followed closely the NIHR best practice standards for public involvement: inclusive opportunities, working together, support and learning, governance, communication, and impact.

We followed the National Institute for Health and Care Research (NIHR) best practice standards for public involvement closely, especially around inclusive opportunities, working together, support and learning, governance, communication, and impact [29]. The guidance emphasises that PPI should entail ‘active partnership’ with members of the public, including young people or older people and family members or carers with lived experience of a health condition, users of health and social care services, and staff from organisations that represent and support them. This aligns with the moral and ethical principles that individuals have a right to have a say in research that affects them and has been shown to result in improved study enrolment [30].

### Stage 1: Planning

The Sleep Detectives PPI lead (JC) has more than eight years’ experience of working across multiple projects and building PPI infrastructure within the NIHR School for Primary Care Research. This includes relationship-building with charities, health and social care organisations, patients, carers, and parents of young children with specific health conditions. Our initial step was to establish a lived experience advisory panel (LEAP) to guide us through the research study, and to plan and help deliver two hands-on PPI workshops.

### Stage 2: Event – PPI workshops

The co-designed workshops, held at the University of Bristol in May and June 2024, were intended to gain informal feedback directly from children and young people with ND-CNVs within the target age range of 6 to 21 years old, as well as their unaffected siblings and parents, on the use of sleep monitoring devices, early designs of cognitive tasks, and study protocol. Throughout the workshops, attendees had the opportunity to share their views through both verbal and written feedback. Families attending the first workshop (in May 2024) also completed an anonymous online survey in MS Forms about their experiences, and this was shared and discussed in the final session on Day 2 and helped shape how the second workshop (in June 2024) was delivered. Families attending the second workshop (June 2024) were invited to share feedback verbally and afterwards by email.

### Stage 3: Evaluation and reflection

Besides giving feedback on the study design and materials, families and charity representatives supporting them also gave feedback on their workshop experience. Further informal verbal and written feedback were gained from LEAP members and the research team about the whole PPI process. Their feedback was gathered using multiple formats including one-to-one and group discussions and email. This range of approaches enabled flexible input from families, accommodating differences in availability, literacy, language, and comfort with sharing personal views in front of others. The research team were also surveyed on their experiences of the PPI process.

## Results

The results of our PPI activities are presented in three sections: **Outputs of PPI planning** by LEAP members and the hands-on PPI workshops; **Integrating PPI feedback into study design and implementation**; and **Learning from PPI and co-design – reflections from families, charities and researchers.** Family members affected by ND-CNVs are quoted directly but anonymously.

### Outputs of PPI planning

#### LEAP and workshop co-design

Nine parents/caregivers joined the LEAP, along with charity partners from Max Appeal and Unique and members of the Sleep Detectives research team. We conducted online LEAP meetings via Zoom in accordance with best practice in PPI.

In planning the two in-person hands-on PPI workshops, the LEAP gave special consideration to working with children with diverse emotional and behavioural support needs and their parents and followed closely the guidance from McPin Foundation on working with young people [32]. This included ensuring that all costs of travel and accommodation for families were covered, and providing advance information about the venue, personnel, and workshop activities, so that families felt fully supported and prepared. The LEAP developed materials for inviting and preparing families for attending the workshops. These included an information pack (with a floor plan, social story, and a timetable of weekend events), and videos that children could view in advance to help them know what to expect and to introduce the research team (see **Supplementary Materials** for links to: **Additional File 1** with the information pack sent to families in advance of attending the PPI workshops; **Additional File 2** showing a video sent in advance to families to explain the iPad cognitive tasks that children and young people will be invited to try out at the PPI workshops; **Additional File 3** showing a video animation to explain the science behind the study.)

The LEAP also gave input on the design of iPad-based cognitive tasks that children would be asked to try out at the workshops. The LEAP’s guidance was critical to ensure that the workshops catered to children’s sensory and emotional needs, see **Table 1**.

**Table 1:**
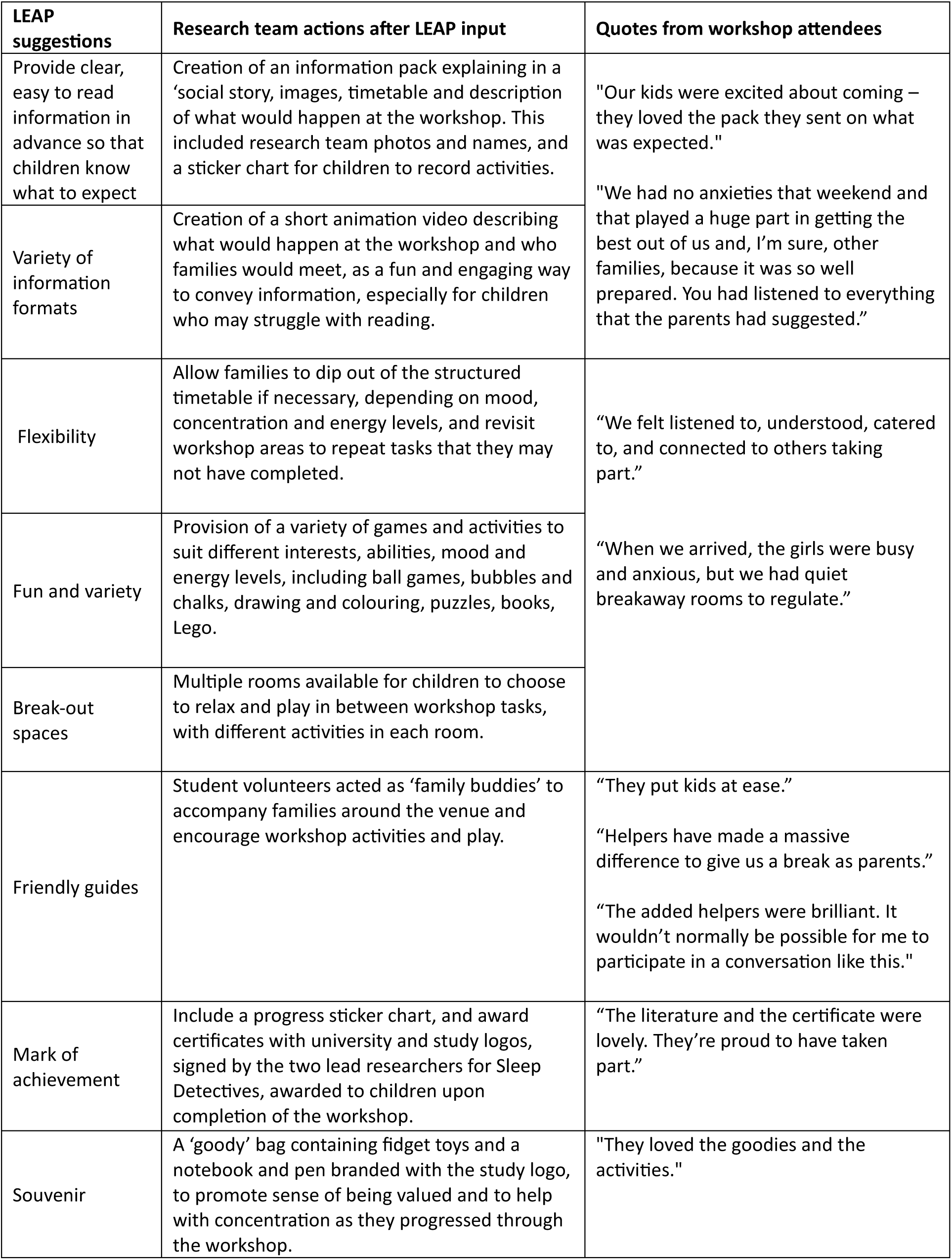
Lived experience advisory panel recommendations for family workshop preparations and impact.

#### Event – PPI workshops

The workshops each took place over a weekend, with two nights’ accommodation provided, to maximise the chance that families could attend without having to take children out of school or for parents and carers to have to take annual leave. The workshops were hosted in a University of Bristol building (Canynge Hall), which has multiple rooms in which to accommodate a flexible workshop environment and timetable (**Figure 2**).

**Figure 2.**
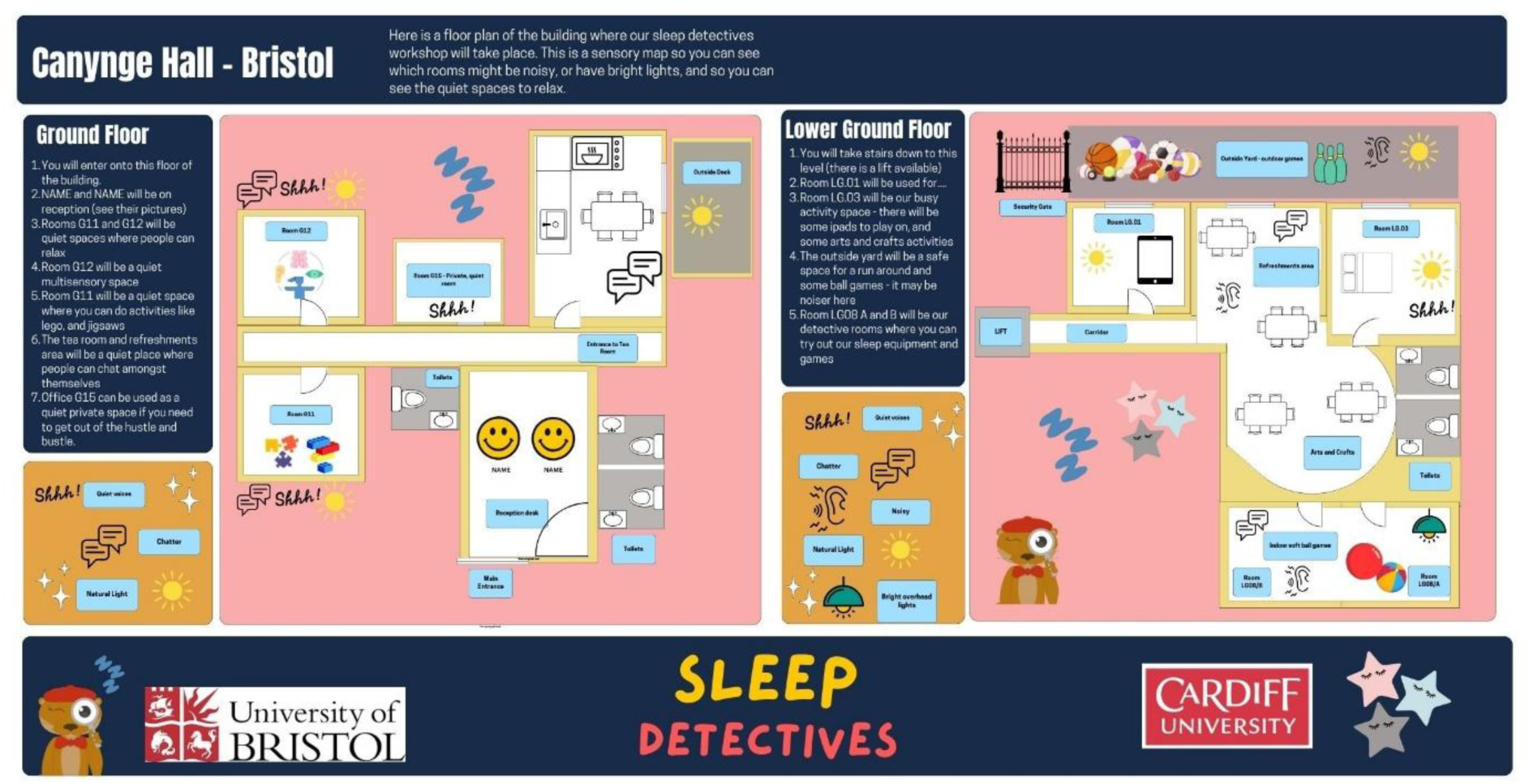
Sensory map showing workshop floor plan Legend. The Sleep Detectives Lived Experience Advisory Panel (LEAP) advised on the need to provide an information pack for children and young people who would be attending the hands-on PPI workshops, including a sensory map showing the layout of the workshop venue and what they would see, hear and do in each room.

In total, 18 families and three charity representatives from Max Appeal and Unique, attended across the two workshops, to try out and give feedback on sleep devices and iPad cognitive tasks (**Figure 3**, **Figure 4 and Figure 5**). The families included 30 children and young people (aged 7 to 19 years old) affected by the ND-CNVs that the Sleep Detectives study is focussing on, including 22q11.2 deletion, 16p11.2 duplication, and 1q21.1 deletion. Families were supported by a total of 26 staff, students and other volunteers from Bristol and Cardiff Universities plus one school A Level student.

**Figure 3.**
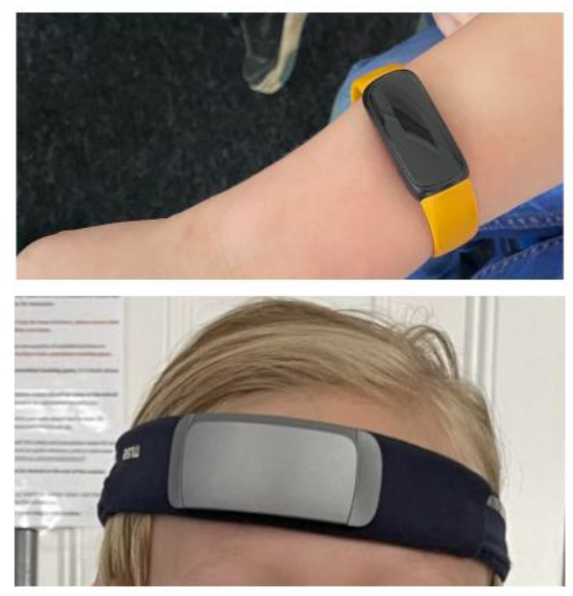
Muse EEG headband and Fitbit actigraphy watch Legend. Children and young people attending the hands-on PPI workshops tried on a range of EEG recording devices including the Muse headband (bottom) and the smaller stick-on Huru Lab Clic EEG sensor, and Fitbit actigraphy watch (top), to give feedback to researchers on their acceptability and ease of use.

**Figure 4.**
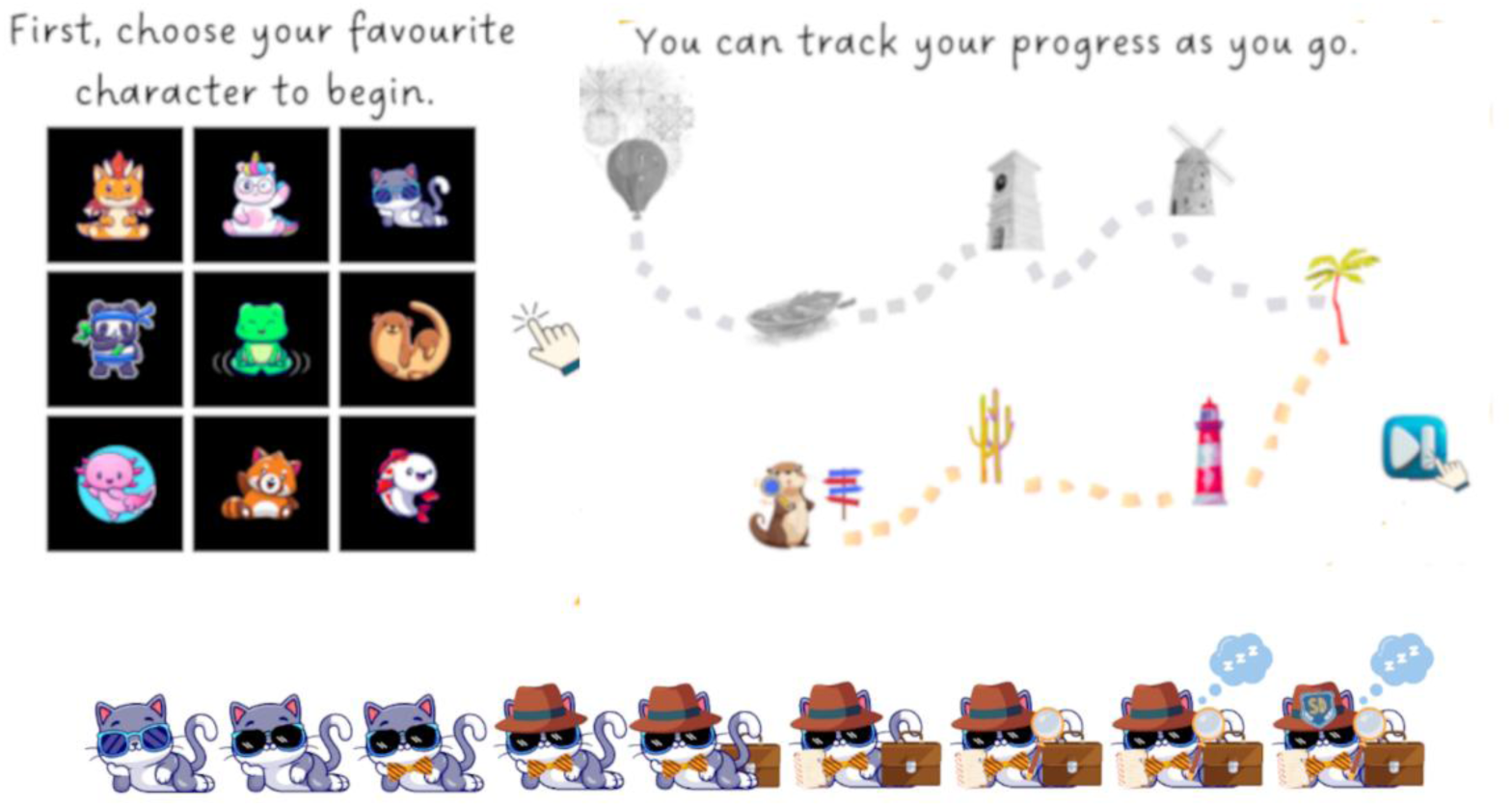
Designs for avatars and tracking of progress through cognitive tasks Legend. Cognitive tasks were designed to appeal to children and young people with friendly-looking and colourful characters, with options to choose an avatar, track progress through tasks that study participants would undertake daily over 7 to 14 days in the main study, and opportunity to trade rewards (gems) for detective gear, e.g., hat, briefcase, magnifying glass and a badge, as tasks are completed.

**Figure 5.**
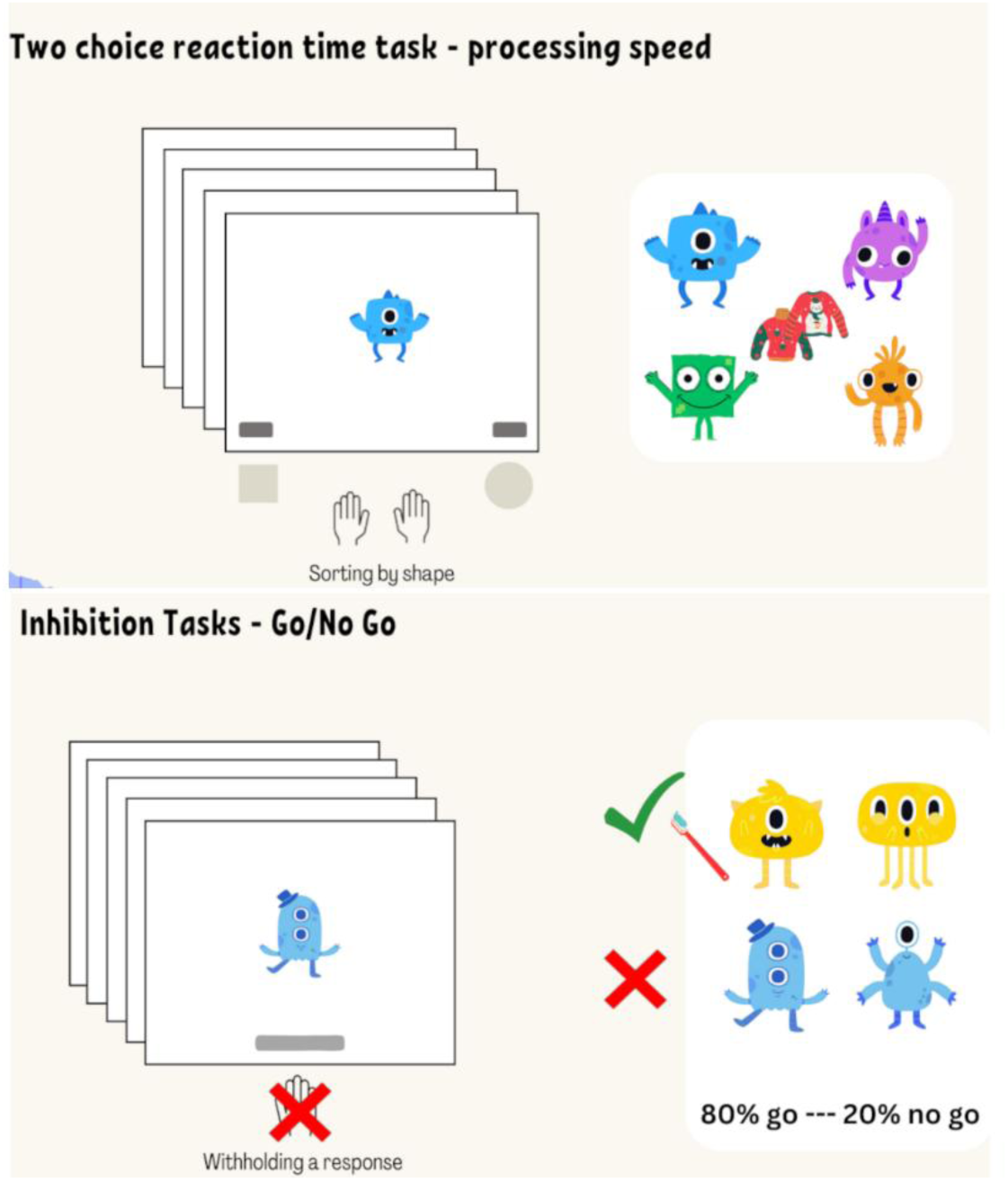
Designs for individual cognitive tasks Legend. Examples of designs for iPad-based cognitive tasks to assess a range of skills including processing speed, inhibition, working memory, and sustained attention.

The feedback from families attending the first workshop (May 2024) provided especially helpful information on what they had enjoyed and recommendations for improvements for the second workshop (June 2024); it also informed plans for the main study.

#### Integrating PPI feedback into study design and implementation

From the LEAP meetings and the workshops, the research team received verbal and written feedback and reflections on the overall study design and protocol, sleep devices, and iPad cognitive tasks (summarised in **Table 2a**, **Table 2b and Table 2c).** This ranged from individual preferences for different sleep monitoring devices, to discussion of the challenges families may have in participating in the study, and refinements (e.g., audio description) for the cognitive tasks. In the survey, 100% of respondents (15/15) were positive (53%) or very positive (47%) about the overall study, and 80% (12/15) thought the protocol would be feasible for their child. Feedback on the sleep EEG devices was more varied. Children and young people were evenly divided between preference for Muse S (Interaxon, Canada) headbands (no sticky gel required) or smaller Huru Clic EEG forehead stick-on sensor. Only 53% (8/15) thought the headband would be comfortable or very comfortable, although most families (80%) were positive about using actigraphy devices.

**Table 2a:** Impact of LEAP and workshop feedback on overall study design and information.

| **Aspect of study design** | **Workshop and/or LEAP feedback** | **Impact on study design** |
| --- | --- | --- |
| Information about the Sleep Detectives study | A video introducing the study would help with understanding and decision-making prior to consenting to take part in the study. 100% of survey respondents (15) were positive (53%) or very positive (47%) about the overall study. 80% (12/15) thought the protocol was feasible for their child. | Development of two information videos for potential study participants: a short (1 minute) and longer (2 minutes and 30 seconds) version. |
| Study information sheets and documents | Keep information simple – bear in mind that parents/caregivers may also have the same genetic condition. | More simplified language in full length participant information sheets and simple-to-read versions created with illustrations. Social story created for children and a sleep calendar with which to monitor progress throughout the study. |
| Timing of tasks at home | Verbal concern about tight morning routine making it tricky to fit in and complete all iPad cognitive tasks. 80% of survey respondents thought evening iPad tasks were achievable, but only 40% thought this of morning tasks. | Reduced length of planned morning session to 5 minutes to maximise engagement and kept evening session to 10 minutes. |
|  | Need for choice for participation in either term time or school holidays when family routines may be different. | Adapted protocol to allow flexibility in data collection; increased the data collection window to allow practice and breaks. Families free to decide when to complete sleep monitoring phase to best fit with schedules. |
| Additional documentation | Write an information letter for schools about wearing the Fitbit and the study in general. | Schools letter included in study information and documentation. Provision of school absence letters for children who may miss school due to study participation. |
| Access to participant data | Request to have a participant's data shared with parents/caregivers including about sleep, cognition, mental health and genetic information. For many parents, receiving this would be a stronger motivation to participate than monetary compensation. | Team agreed to provide individualised reports on cognitive, sleep and neuropsychiatric assessment where possible. In line with current practice for non-clinical (research) genomic analysis, genomic data will not be shared. |
| Feeding back study updates and results | Suggestion that a general summary newsletter to participants /all interested parties might be useful. | Developed a bi-annual newsletter and a website blog, to provide study updates. |

**Table 2b.** Impact of LEAP and workshop feedback on sleep devices and protocol.

| **Sleep devices / protocol** | **Workshop and/or LEAP feedback** | **Impact on study design** |
| --- | --- | --- |
| EEG devices | Children and young people were evenly divided between preference for Muse headbands (no sticky gel required) or smaller Huru Clic EEG forehead stick-on sensor. Some said they did not like the sticky sensation of the forehead sensor. Others said this was acceptable especially if the sensor could be customised with a colourful or cartoon image sticker. Some were not keen on the sensation of tightness or itchiness with the headband. Only 53% (8/15) said they would be comfortable or very comfortable wearing the sleep headband for 7 nights. | Researchers recognised that unfortunately no single device suited all; they chose the device that, with further testing, produced the most consistent and reliable data; they explained the decision to LEAP members. |
| Actigraphy watch (Fitbit Inspire 3) | Children found these comfortable. But some suggested that wearing this during the day at school may feel embarrassing to be conspicuous among peers. 80% of survey respondents (12/15) said their children would be comfortable or very comfortable wearing the actigraphy watch. Children would love the wristbands to be branded with the Sleep Detectives logo. | Created customised sweatbands with the Sleep Detectives mascot for children to place over the actigraphy watch during school physical activities. The sleep kit has been adapted to include different fabric straps to allow for both choice and sensitivities to different materials. |
| Sleep monitoring protocol | Families said they would be willing to try the sleep gear at home, and that 7 nights' continuous monitoring was reasonable. But they cautioned that some issues might interfere and so limit data collection, including disruption if children divided time between different houses, as well as mood and behaviour changes. Children may therefore need a break of one or more nights in between data collection nights. | Adapted the protocol to allow for flexibility, increasing the data collection window to allow for practice and breaks. Families will also be able to decide when they would like to complete the sleep monitoring phase of the study to best fit with their schedules. |
|  | Verbal feedback that children should have one or more practice nights before Day 1 of testing, so that they can get used to the sleep wearables. 93% of survey respondents (12/15) thought a practice night would be useful. | Protocol includes an optional practice night with the sleep monitoring devices. |

**Table 2c.**
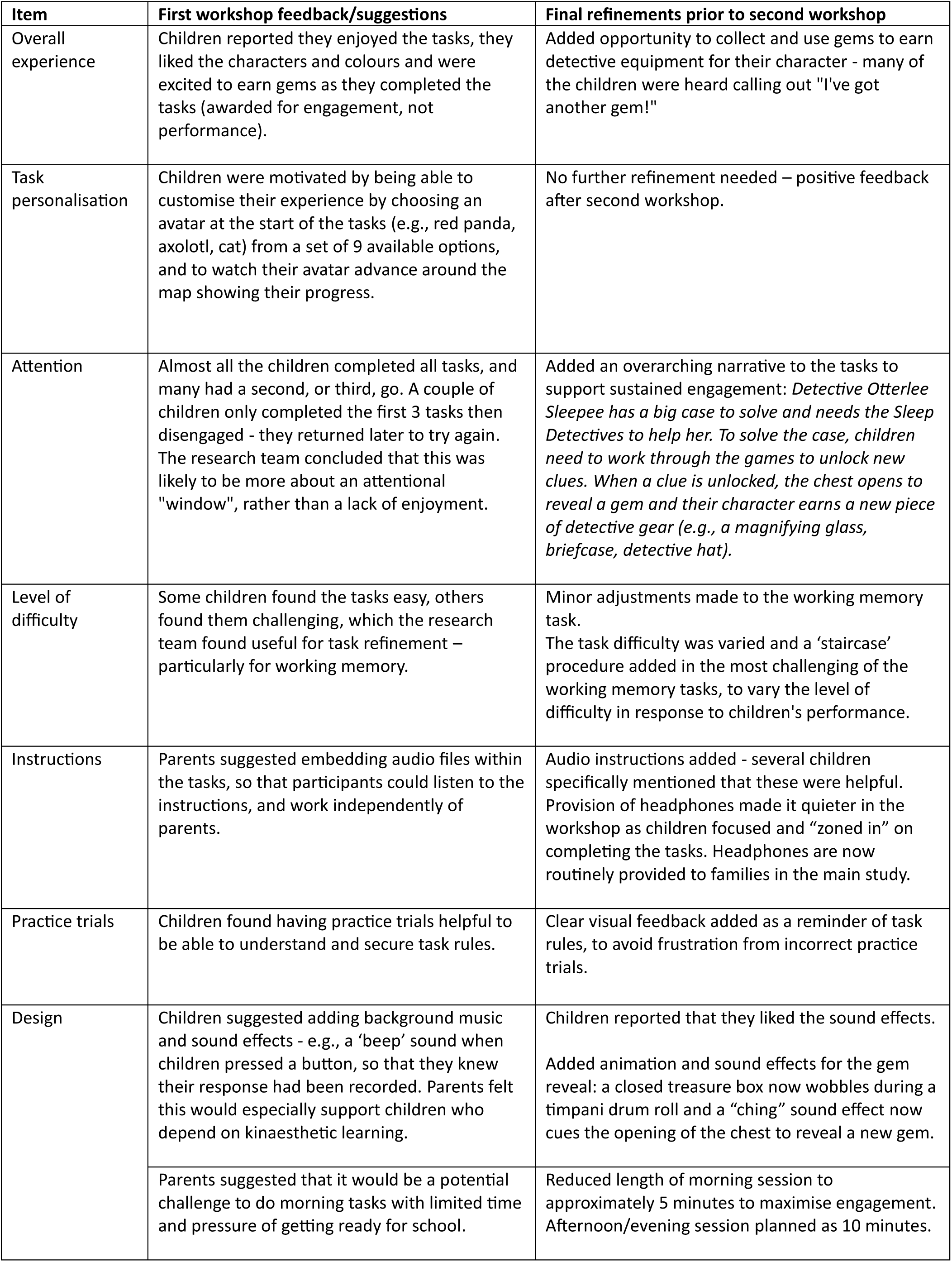
Impact of PPI on cognitive task design and refinements between workshops.

| Item | First workshop feedback/suggestions | Final refinements prior to second workshop |
| --- | --- | --- |
| Overall experience | Children reported they enjoyed the tasks, they liked the characters and colours and were excited to earn gems as they completed the tasks (awarded for engagement, not performance). | Added opportunity to collect and use gems to earn detective equipment for their character - many of the children were heard calling out "I've got another gem!" |
| Task personalisation | Children were motivated by being able to customise their experience by choosing an avatar at the start of the tasks (e.g., red panda, axolotl, cat) from a set of 9 available options, and to watch their avatar advance around the map showing their progress. | No further refinement needed – positive feedback after second workshop. |
| Attention | Almost all the children completed all tasks, and many had a second, or third, go. A couple of children only completed the first 3 tasks then disengaged - they returned later to try again. The research team concluded that this was likely to be more about an attentional "window", rather than a lack of enjoyment. | Added an overarching narrative to the tasks to support sustained engagement: *Detective Otterlee Sleepie has a big case to solve and needs the Sleep Detectives to help her. To solve the case, children need to work through the games to unlock new clues. When a clue is unlocked, the chest opens to reveal a gem and their character earns a new piece of detective gear (e.g., a magnifying glass, briefcase, detective hat).* |
| Level of difficulty | Some children found the tasks easy, others found them challenging, which the research team found useful for task refinement – particularly for working memory. | Minor adjustments made to the working memory task.<br>The task difficulty was varied and a 'staircase' procedure added in the most challenging of the working memory tasks, to vary the level of difficulty in response to children's performance. |
| Instructions | Parents suggested embedding audio files within the tasks, so that participants could listen to the instructions, and work independently of parents. | Audio instructions added - several children specifically mentioned that these were helpful. Provision of headphones made it quieter in the workshop as children focused and "zoned in" on completing the tasks. Headphones are now routinely provided to families in the main study. |
| Practice trials | Children found having practice trials helpful to be able to understand and secure task rules. | Clear visual feedback added as a reminder of task rules, to avoid frustration from incorrect practice trials. |
| Design | Children suggested adding background music and sound effects - e.g., a 'beep' sound when children pressed a button, so that they knew their response had been recorded. Parents felt this would especially support children who depend on kinaesthetic learning. | Children reported that they liked the sound effects.<br>Added animation and sound effects for the gem reveal: a closed treasure box now wobbles during a timpani drum roll and a "ching" sound effect now cues the opening of the chest to reveal a new gem. |
|  | Parents suggested that it would be a potential challenge to do morning tasks with limited time and pressure of getting ready for school. | Reduced length of morning session to approximately 5 minutes to maximise engagement. Afternoon/evening session planned as 10 minutes. |

### Overall impact

The involvement of families affected by ND-CNVs and the encouragement and support provided to them by charity partners, had significant impact on the study protocol and materials. Moreover, the research team gained insight into the preferences and opinions of families with lived experience of ND-CNVs. They listened to families and implemented their recommendations appropriately. While there was diversity in views, the process allowed the study team to improve the study design by implementing several changes which are summarised below.

#### Impact 1: study participation – the need for flexibility

Discussions about the burden on families, who are already “time-poor”, highlighted the importance of a study design which minimised the time required by parents and caregivers to supervise involvement and maximised understanding and acceptability for many of these children and young people with ND-CNVs. Many will exhibit challenging behaviours and struggle with everyday activities such as getting ready for school. Difficulties with changes in schedules or sensory sensitivities are also common. Changes made to the study design included reducing the duration of the cognitive tasks and allowing some of the tasks to be completed at a time decided by the participants. The study team also increased the data collection window from eight days to two weeks to allow participants to take breaks between the 8 days/nights of data collection when needed. They also added the opportunity for practice nights with the sleep monitoring equipment. In a further response to workshop feedback the team agreed to include provision of school absence letters for children who may miss some school due to participation in the research.

#### Impact 2: communication with participants – multimodal delivery

LEAP members highlighted the importance of giving study information and supporting documents to the parents/caregivers and children and young people taking part in easy to understand and developmentally appropriate formats. The LEAP emphasised that research information can often be confusing and should be simply worded. The LEAP and research team co-designed workshop materials including information sheets, social stories, sensory maps, and photographs of the researchers and volunteers who would be attending the events, as well as branding (for example, the Sleep Detectives mascot, Detective Otterlee Sleepee), to encourage recruitment of families to the workshops. This PPI work provided the foundation for the subsequent approach and design of information sheets and recruitment materials for the Sleep Detectives research study. Accordingly, in collaboration with LEAP, the Sleep Detectives research team have produced two introductory videos for the study, simplified versions of all recruitment documents, and a website with study information in an easy to digest manner (https://sleepdetectives.blogs.bristol.ac.uk) [33]. These relatively inexpensive information resources are expected to have a significant impact on study recruitment and thus on achieving the main research aims. The team also developed a bi-annual newsletter to provide updates and progress on the study to participating families.

#### Impact 3: selection of sleep recording devices

Families appeared to be evenly split in their preference of the two types of brain EEG measuring devices: a headband (Muse S, Interaxon) and one that sticks onto the forehead (Clic EEG, Huru); there was no single device that perfectly suited all workshop participants (see **Figure 3** showing the Muse headband and actigraphy device (Fitbit Inspire 3)). Following subsequent pilot-testing and evaluation of the different EEG devices, the research team concluded that the Clic EEG device provided more consistent and high-quality data. They explained to LEAP members in meetings and by email that this device was ultimately selected for consistency, despite some children at the workshops having expressed a preference for the headband. The research team acknowledged that this may have a potentially negative impact on study recruitment and retention if some participants were less keen on the Clic EEG device, but that the improved data quality and consistency was necessary to complete the research aims. LEAP members accepted that this was a necessary decision. Children requested to be able to customise the EEG headbands or stick-on devices, with a choice of stickers displaying different characters. The Clic EEG developers (Huru) made this possible, and the study currently has electrode stickers customised with the Sleep Detectives mascot, Detective Otterlee Sleepee. Many families were very positive about the use of nearable devices (placed close to the person sleeping) to monitor sleep, and therefore the research team investigated several different devices including under-the bed and bedside devices. The study now includes a bedside device (Somnofy, Vitalthings) as part of the sleep measuring kits.

The feedback was also positive about the use of actigraphy watches (Inspire 3, Fitbit), but some concerns were raised about comfort and use during physical activity. In response, the study sleep kits include different options for actigraphy watch straps – both fabric and rubber - to allow for different sensory preferences. For the main study, we have also included a Detective Otterlee Sleepee-branded towelling wristband (e.g. to cover the actigraphy watch during P.E.), that the participants will be able to keep. These adjustments aim to ensure more continuous data collection during the study period.

#### Impact 4: improving accessibility of the cognitive tasks

The workshops provided useful feedback on the cognitive tasks (see **Figure 4 and Figure 5** showing designs for tracking progress and individual cognitive tasks). In addition to flexibility/timing of tasks, additional feedback from the first workshop included suggestions for the addition of audio instructions and headphones with the sleep kits, and the customisation and gamification of the cognitive tasks. These changes were made before the second workshop and were positively received by families. Children and young people liked the changed characters and colours and were excited to earn gems as they completed each task. Importantly, these gems are given on completion of each task to reward engagement regardless of how the individual performed on that task, and so are not dependent on levels of performance.

#### Learning from PPI and co-design – reflections from families, charities and researchers

### The importance of studying sleep: lived experience

#### Box 1: Years of poor sleep, affecting the whole family

Families have described to the Sleep Detectives team the sleep problems experienced by children and young people with Copy Number Variants. M, for example, now adult, has struggled since childhood to get a good night’s sleep. “It’s not great. I wake up in the night. I have night terrors. I sometimes come down the stairs. I disrupt everyone. I just don’t have very good sleep at all. When I wake up in the morning and I feel quite grumpy, it just ruins the rest of my day.”

M’s mother says: “It affects both of us - that’s the problem. I don’t get good sleep, and I haven’t had for years.”

Similarly, A tells us that her daughters had problems sleeping from birth. Initially she thought this was due to having a two and a half-hourly feeding schedule in the hospital neonatal unit where they went straight after birth. “After I brought them home, for years they woke up at the same hours that they were fed. Eventually this changed but sleep remained difficult: settling to sleep, falling asleep is extremely difficult. Sometimes it takes two hours. And then staying asleep is also very difficult.”

The resulting tiredness can cause significant issues at school. L says about her son: “Sometimes school will comment and put in his care diary that he has been very tired, sleepy and moody. He does sometimes have a nap in his taxi on way there and on way home as well, he often arrives at school having just woke up.”

For parents too, the impacts can be considerable. A says that despite following all the guidance on bedtime routines for his now teenage daughter, “It affects the family. It means you can’t concentrate if you’ve got a job to hold down, you’re falling out with your loved ones, it causes relationship breakdown without sleep.”

The relevance of studying sleep in children and young people with ND-CNVs was reinforced by parents/caregivers of children with ND-CNVs, and by charity partners from Max Appeal and Unique. **Box 1** summarises some of these experiences and highlights the impact of disrupted sleep on the whole family.

### Wider impact: families and researchers of the PPI experience

These reflections were gathered to illuminate the broader value of patient and public involvement, demonstrating not only its contribution to effective study design, but also its personal and relational impact on all the stakeholders involved, including feelings of connection, empowerment, and shared purpose.

Feedback and reflections from LEAP parents and charity representatives and researchers on their PPI experiences are provided in **Table 3**. This feedback was collected for the purpose of gaining insight and improving the PPI process in future. The feedback was not subject to thematic analysis or other qualitative research methods. The research team heard positive feedback from workshop families and LEAP members about their enjoyment of being involved in a research project – for most families this was their first experience of research. Families reported that they felt listened to, and that their suggestions contributed to significant decisions about study design. In agreement with previous findings, we found that the lived experience advisors on Sleep Detectives gained a new understanding of research and a renewed sense of purpose [34]. Workshop attendees also made new friendships with other families and found peer support, including setting up a WhatsApp group. LEAP members felt encouraged to seek new opportunities, including appointment to a new forum/taskforce, and have opted to continue as lived experience advisors on other research studies.

**Table 3:**
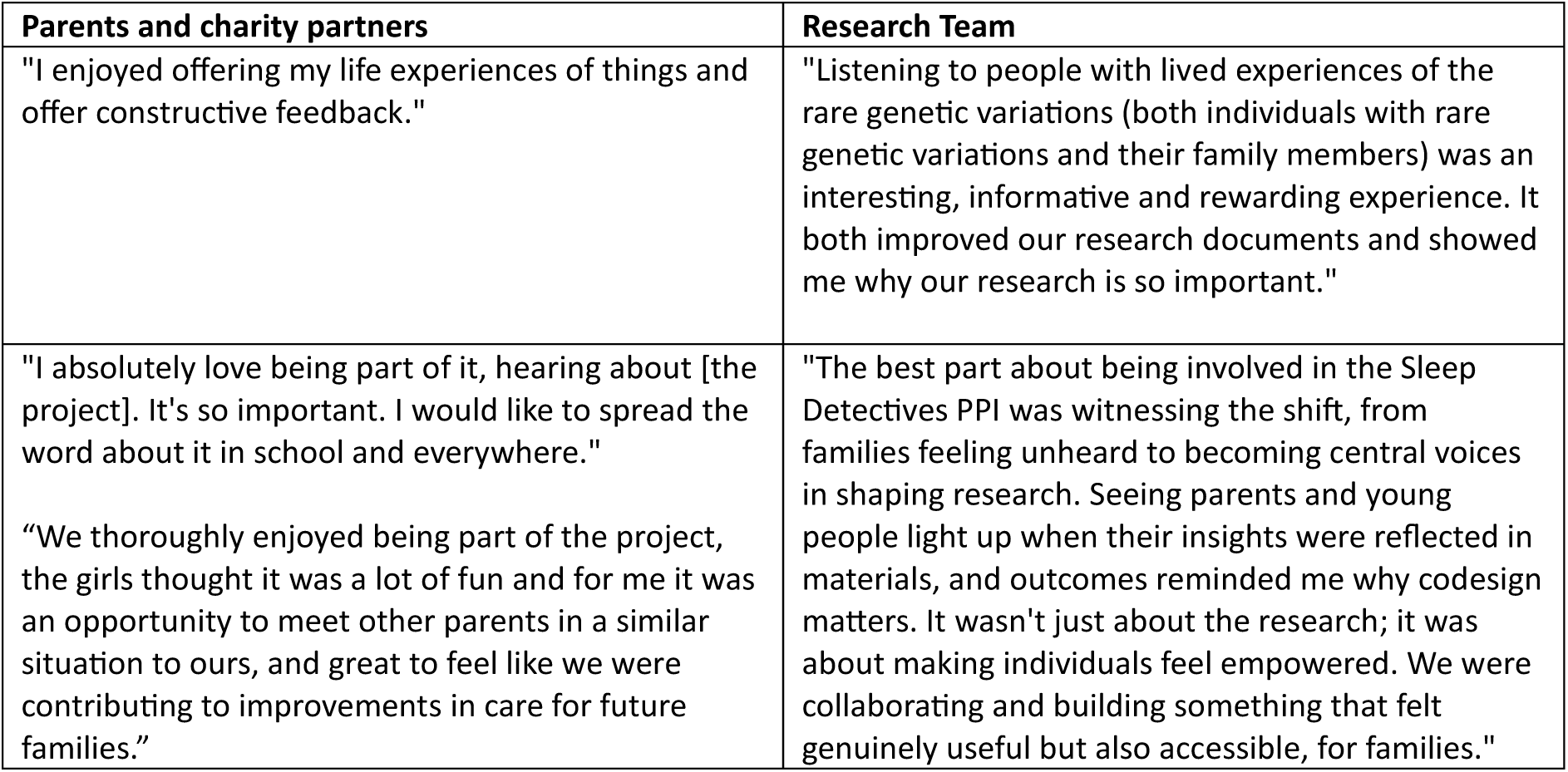
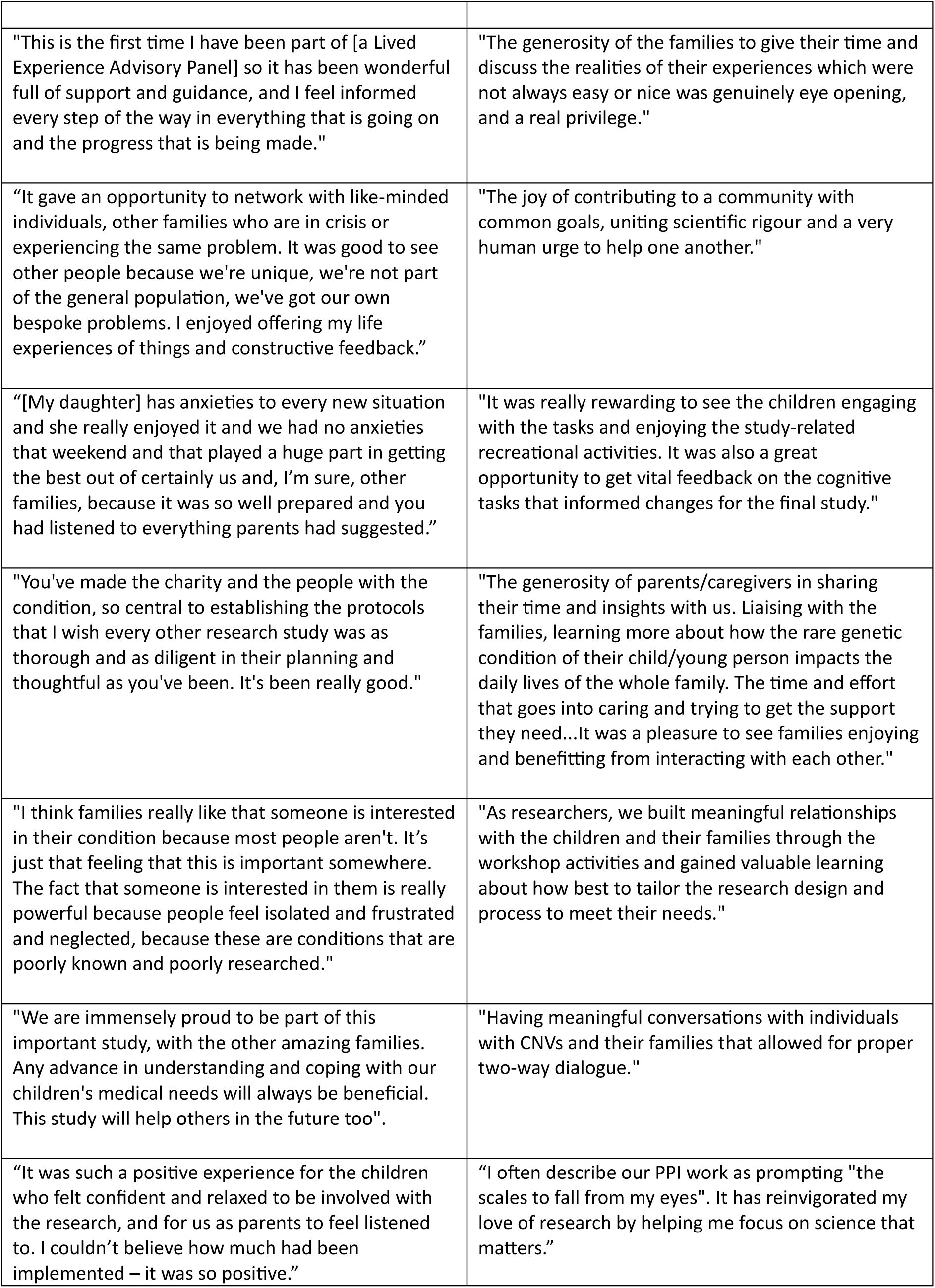
Reflections from LEAP members on the PPI process.

Some of the Sleep Detectives study team had a background in laboratory-based neuroscience and psychology. They had not previously worked with families affected by ND-CNVs, or had much PPI experience, but were well prepared for conducting the workshops, thanks to guidance from parents and charity partners on the LEAP. 100% of the Sleep Detectives research team felt that the workshops and LEAP meetings were very useful. Challenges included scheduling of meetings to meet everyone’s availability and the cost to the research team of doing weekend workshops, the emotional load (i.e. the mental and emotional effort required to manage feelings - both those of the research team and LEAP members) especially when realising first-hand how many challenges these families face, navigating sensitive topics, and how to deal with differences of opinion especially when the research team were unable to meet some of the requests of families. Positives included the refinements to study design which will improve the integrity and impact of the research, and the inspiration felt by working together. All the research team reported wanting to continue to do PPI work in the future.

## Discussion

### ‘You said, we did’

The Sleep Detectives team have conducted extensive PPI which has formed the foundation for development, refinement, and testing of the study’s equipment and cognitive tasks, recruitment materials, and protocol. Children and families with lived experience of ND-CNVs played an integral role in shaping the Sleep Detectives study protocol and have actively contributed to decision-making regarding data collection methods and equipment, to ensure that each component was accessible, relevant, and finalised in response to the needs and experiences of families and children with ND-CNVs. Recruitment materials and ethics documents have been co-created with the LEAP ensuring that information is appropriately tailored and understandable, and that consenting and data collection processes are acceptable and feasible for participating families.

During the workshops and LEAP meetings, parents/carers, children and young people were able to share the challenges they face and the impact these have on their everyday lives. Their lived experience has informed the research team’s understanding of the daily impact of ND-CNVs on family life and highlighted issues that could affect recruitment, compliance, and data quality in the main study. Early and ongoing opportunities to share feedback on the sleep monitoring equipment (e.g., choice of EEG devices) and the cognitive tasks have afforded families the opportunity to play an active and meaningful role in co-designing these aspects of the study.

The ability to influence the study design brought positive feelings of engagement and ownership for families, and of having their voices heard. Through PPI, parents and caregivers had the opportunity to link up with charity representatives, meet other families, and form supportive peer networks that run independently of the Sleep Detectives study that will be sustained beyond families’ involvement in the project. For researchers on the Sleep Detectives team, in addition to improving “essentially everything” (quote study PI) in the study design, and recruitment and engagement processes, the PPI work was inspiring, motivating, and a driving force to do more accessible and relevant research in the future.

There were several challenges along the way. Discussions around the difficulties and fears faced by families of children with ND-CNVs carried a significant emotional weight, for both those generously sharing their experiences and the researchers listening. This was particularly evident in moments where researchers were unable to offer direct support or solutions. Another key challenge was how to navigate tensions between the diverse needs and preferences of ND-CNV families and the methodological or logistical requirements of the research. The research team learned, from the experience of establishing a LEAP and running the family workshops, that it would not be possible to achieve consensus in all discussions – for example, regarding the choice of brain EEG recording device; balancing these perspectives required ongoing dialogue, flexibility, and a commitment to maintaining trust while ensuring scientific rigour.

It was important for all voices to be heard, and for researchers to be transparent about the justification for each decision, explaining the limits to what could be changed following feedback. It was crucial to be prepared from the start for the challenge of balancing divergent opinions, and how potential differences would be resolved [35]. For example, it was discussed at the initial LEAP meeting that some aspects of the study design may have to be decided by the research team despite LEAP members and workshop participants expressing alternative preferences, but that careful and respectful consideration would be given to all suggestions, and the justification for any contrary decisions would be explained clearly. LEAP members were generally positive about this process, with one quote reflective of this: “I feel informed every step of the way in everything that is going on and the progress that is being made”. Future challenges will be to further widen inclusion and access for both PPI in research design and research participation, to gain a broader representation of families with ND-CNVs, and to use these co-designed foundations to extend to multi-year, longitudinal studies, which are crucial to understand the development of children’s strengths and weaknesses over time and to inform prevention and early intervention strategies in the future.

## Conclusions

It was important to involve families affected by CNVs in co-designing the Sleep Detectives study protocol, and materials: co-design ensures that the study remains grounded in scientific rigour, whilst also ensuring that the protocol is acceptable and not burdensome to participants. This may prove particularly vital for studies dealing with complex aspects such as sleep and mental health and more vulnerable cohorts such as children, and children with CNVs, potentially over multi-year timescales. The LEAP insights helped the research team tailor the study to better meet families’ needs, fostering trust and enabling meaningful participation. As many research studies are negatively affected by poor recruitment and retention, particularly with regards to underrepresented groups, such PPI is crucial for informing how best to support participation in future studies.

This PPI work also had wider benefits; the research team gained a deeper understanding of the impacts of ND-CNVs on families and the barriers they face to participate in research, as well as a clearer idea of the importance of doing meaningful public involvement. Families gained understanding of the research process, and developed new connections and peer support networks, and were enthusiastic about being involved in research in future. Co-design is therefore not just a methodological choice, but a vital component of ethically grounded and impactful research. In the context of the Sleep Detectives project, embedding participant insight throughout ensured the study is not only methodologically robust, but also meaningfully tailored to the very individuals and families it seeks to understand and support.

## Supporting information

Additional file 3 Animation to explain Sleep Detectives for PPI workshop families

Additional file 1 PPI workshops information pack

Additional file 2: Sleep Detectives introduction to iPad cognitive tasks for PPI workshops

## Data Availability

All data produced in the present study are available upon reasonable request to the authors

## Abbreviations

LEAP: Lived Experience Advisory Panel
PPI: Patient and Public Involvement in research
ND-CNVs: neurodevelopmental copy number variants

## Declarations

### Ethics approval and consent to participate

Ethics approval is not applicable for the PPI advisory-only activities described in this report, in accordance with best practice standards of the UK National Institute for Health and Care Research (NIHR) https://www.nihr.ac.uk/reporting-ppi-publications-guidance

### Consent for publication

All authors contributed to the study and approved the final version for publication. The parents and children involved in the Lived Experience Advisory Panel and PPI workshops provided consent for the inclusion of their testimonials and images in this manuscript.

## Author information

### Contributions

JC led on PPI planning and delivery, including recruitment of parents and charity partners for the Lived Experience Advisory Panel (LEAP), facilitation of LEAP meetings, inviting and briefing families prior to workshops, organising workshop schedules and volunteer rotas, and writing the first and final drafts of this paper. All co-authors participated in LEAP meetings, led or supported activities at the PPI workshops (except JEH) and contributed to drafts of this paper. MBMvdB and MWJ are lead investigators on Sleep Detectives and recruited student volunteers to support the workshops. JH designed the cartoon character Detective Otterlee Sleepee, as well as the workshop invitations and information packs, and incorporated LEAP feedback and co-edited this paper. MA and CJ designed the cognitive iPad tasks for the workshops and refined these following PPI discussions and workshops. LB created videos including animation for children and young people to understand the purpose of the Sleep Detectives study and what to expect at the workshops, in consultation with LEAP members. JEH helped gather feedback from LEAP members, and together with workshop feedback, refined the study protocol and recruitment materials, and co-edited the paper. AS participated in LEAP meetings and with setting up and supporting workshop activities and feedback sessions. All co-authors have read and approved the final version.

### Collaboration

We wish to thank public members of the Lived Experience Advisory Panel for collaboration and support on the project: Lisa Cundall, Anita Czekus, Kathy Goodwin, Alexander Huxford, Keith Huxford, Laura Lavery, Gemma Mackew, Samantha Rogers, Mark Tripp (MaxAppeal), Julie Wootton (MaxAppeal), Sarah Wynn (Unique)).

## Acknowledgements

We greatly appreciate the generosity of the families on the Lived Experience Advisory Panel and other families who participated in the hands-on PPI workshops, and the staff and student volunteers who helped with the workshops (Anna Andrieu, Nuttaya Bunmak, Hayah Chaudhry, Mehtaab Chaudhry, Hannah Davies, Anna Donnelly, Ellie Evans, Abi Gass, Ciara Harris, Will Hughes, Josh Hope-Bell, Danielle Le Roux, Finn Schofield, Shreeya Sivakumar, Hannah Thomas, Thomas Wheatley). Thanks also to Owen Meyers for supporting LEAP administrative tasks, and to Daniel Brooks for helping to incorporate LEAP and workshop feedback into refining the study protocol and for creating the study blog site.

## Note

Further information about the Sleep Detectives study, including videos created to explain about the study for children and young people, can be found at the following URL https://sleepdetectives.blogs.bristol.ac.uk/ [last accessed 4 February 2026]

Further information on our charity partners can be found on their respective websites: Max Appeal https://www.maxappeal.org.uk/ and Unique https://rarechromo.org/

## Supplementary materials

- **Additional file 1 (pdf): PPI workshops information pack.** This contains the information co-produced by the Lived Experience Advisory Panel to explain to children and young people what to expect at the hands-on PPI workshops, who they would meet, what activities they would do, a sensory map of the venue and a sticker chart to mark their progress through the workshop. Accessible at Open Science Framework https://osf.io/hkprx/files/zwc96
- **Additional file 2: Sleep Detectives introduction to iPad cognitive tasks for PPI workshops** (video, MP4). This video introduces the new iPad tasks that children and young people will be invited to try out at the PPI workshops, and explains that these tasks are intended to help researchers understand how sleep affects children’s thinking and learning. The video features co-authors Professor Christopher Jarrold and Dr Meg Attwood, who have granted permission for their images to be used in this report. Accessible at Open Science Framework: https://osf.io/hkprx/files/953vh
- **Additional file 3: Animation to explain Sleep Detectives for PPI workshop families** (video, MP4). This animation video explains the idea of measuring brain activity through EEG, and why this may help to understand the link between sleep, mental health and behaviour. Accessible at Open Science Framework: https://osf.io/hkprx/files/nbxav

