## Additional file 1 PPI workshops information pack for "Public involvement and co-design of longitudinal studies of sleep health alongside young people with rare genetic conditions"

### SLEEP DETECTIVES

CAN YOU HELP DETECTIVE OTTERLEE SLEEPEE
AND HER TEAM OF RESEARCHERS?

WE ARE SEEKING A SQUAD OF SNOOZE SLEUTHS TO CRACK THE CASE OF "THE ELUSIVE EQUIPMENT"!

#### WORKSHOP:
#### 8-9TH JUNE

CAN YOU TRY
OUR EQUIPMENT AND
GAMES
AND TELL US
WHAT YOU THINK?

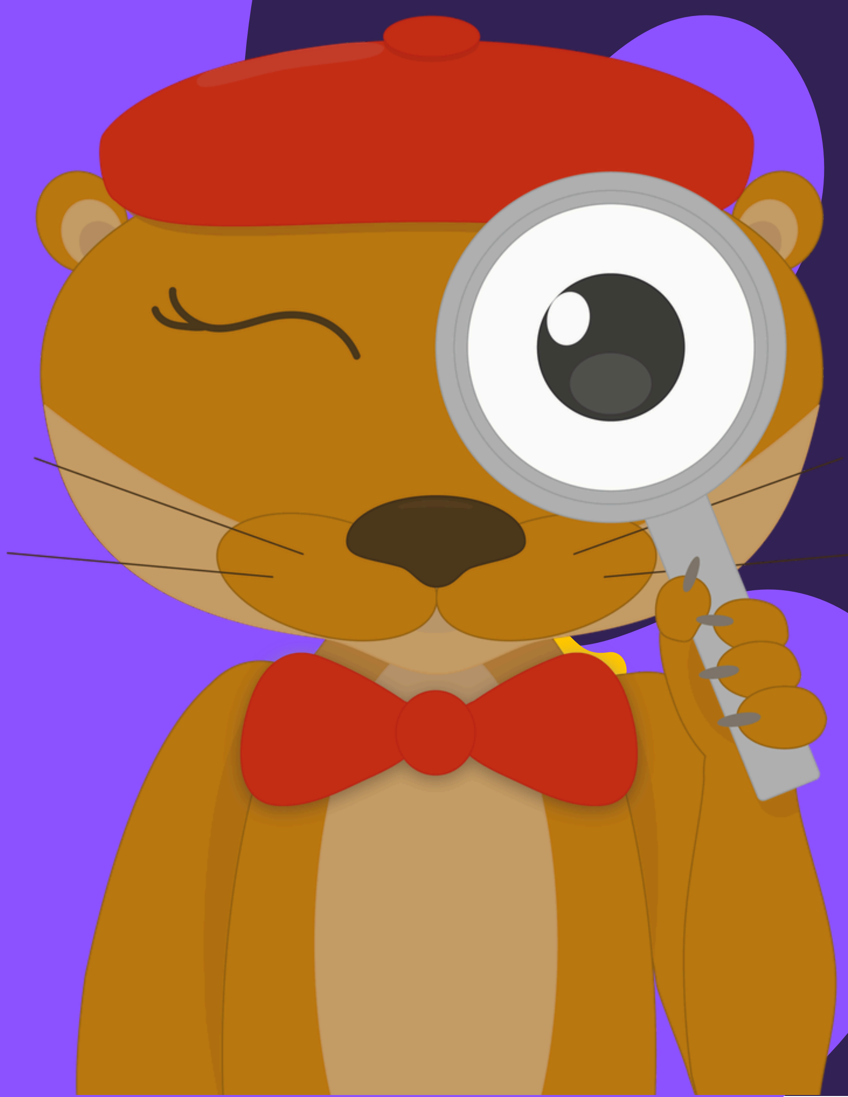

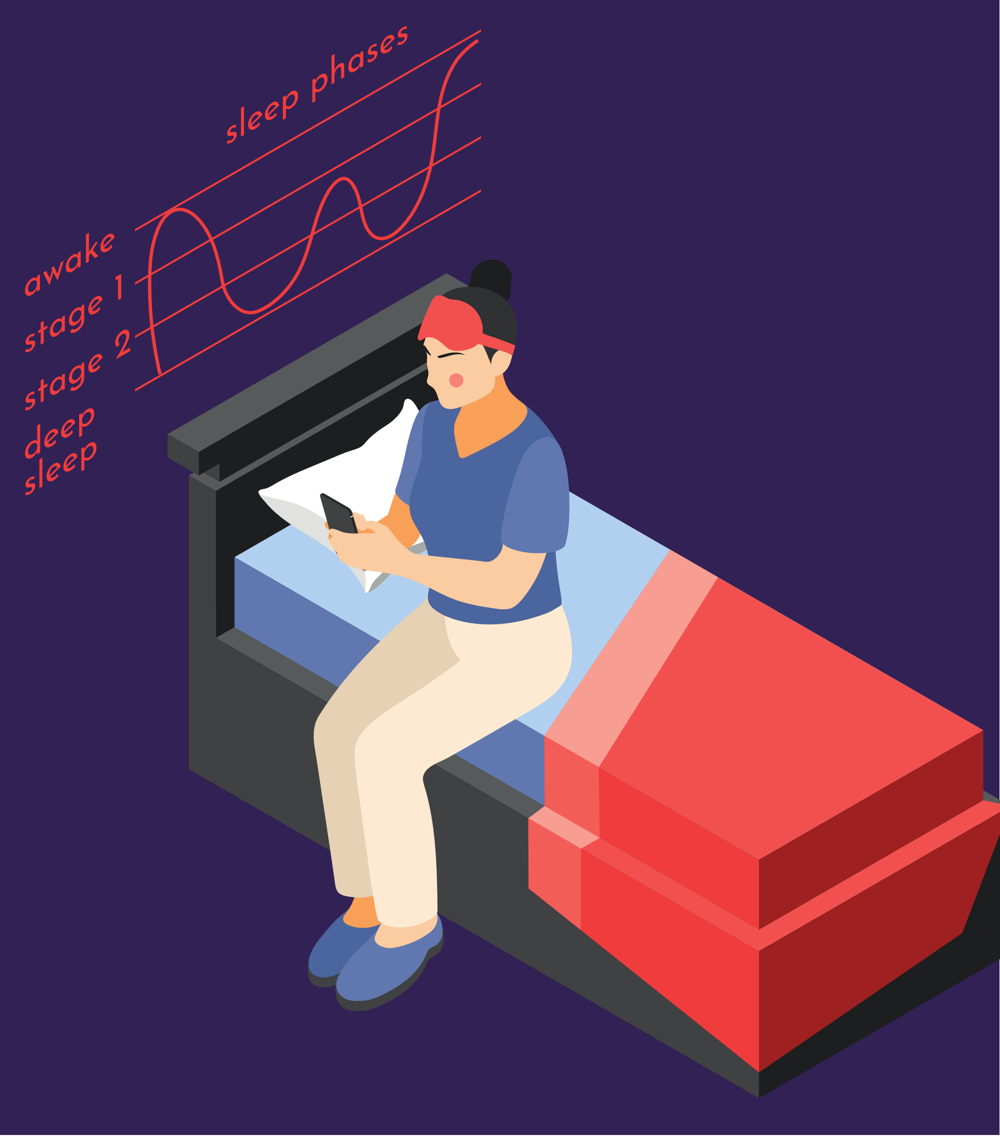


Bring your families:
CANYNGE HALL
Bristol

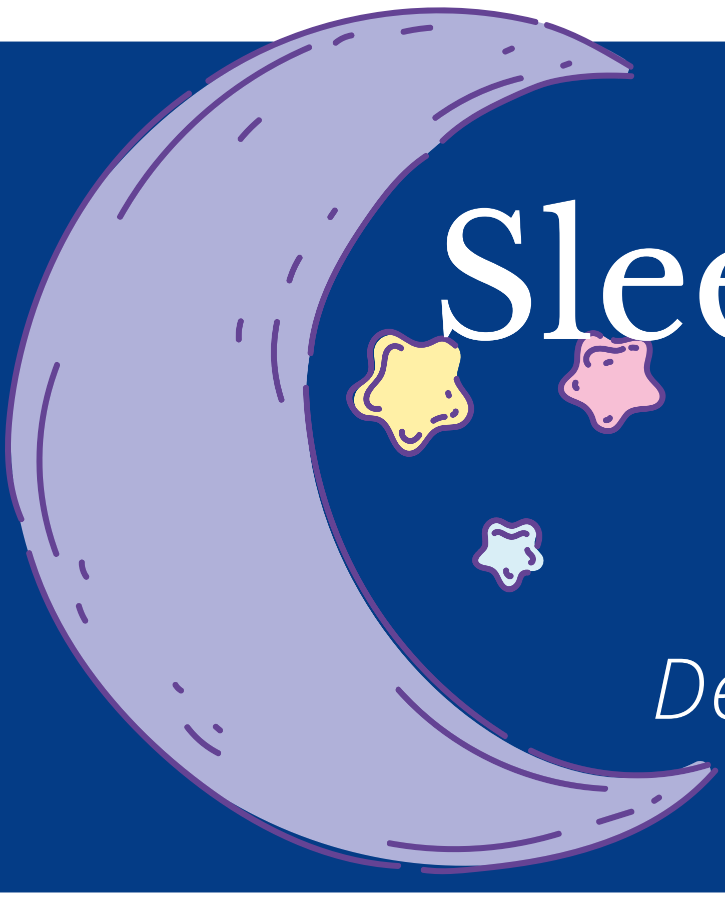

### Sleep Detectives

*Unravelling the secrets of sleep with Detective Otterlee Sleepee*

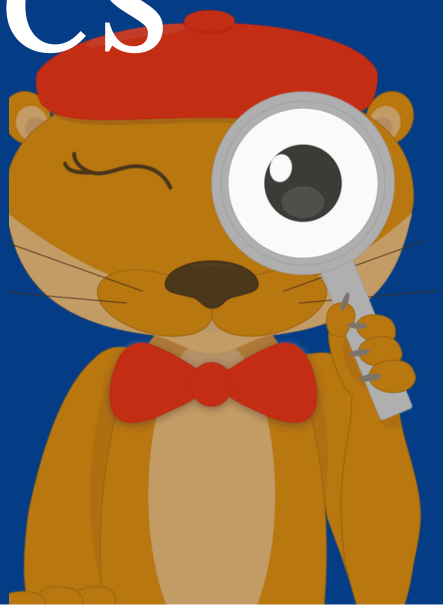

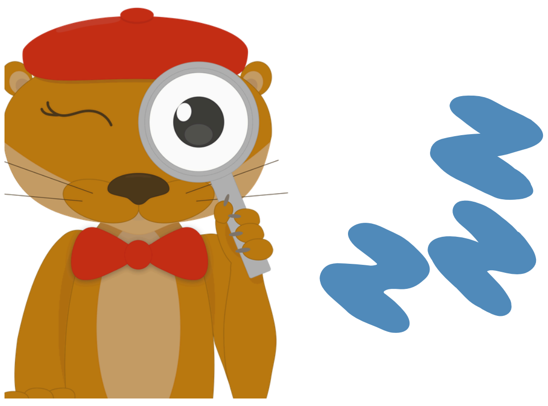

Detective Otterlee Sleepee, and her group of researchers are interested in how different children might sleep differently.

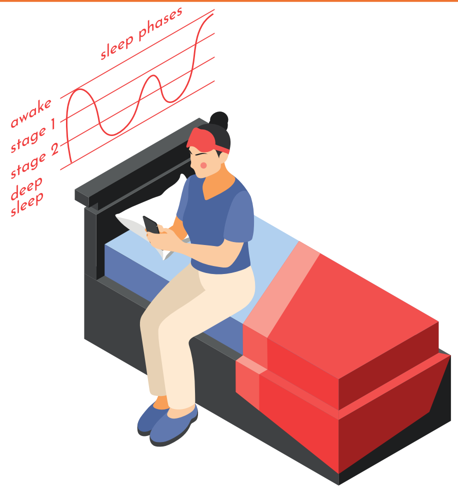


To understand more about how children sleep, researchers need to use special equipment.

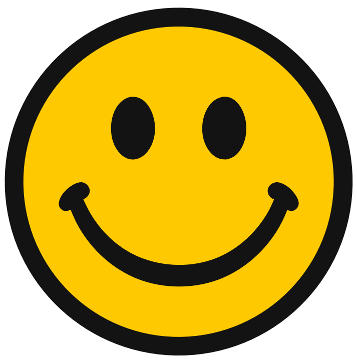

The researchers need to make sure that the equipment they use is the best, and that it is comfy.

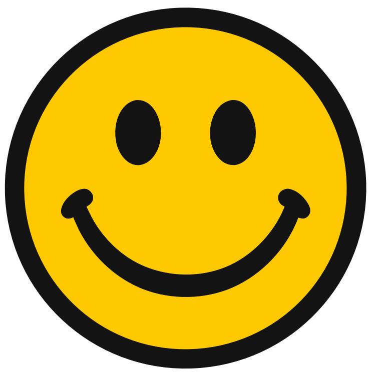

Can you be a sleep detective, and help Detective Otterlee Sleepee and her gang of researchers, by trying out their equipment?

University of BRISTOL

CARDIFF UNIVERSITY

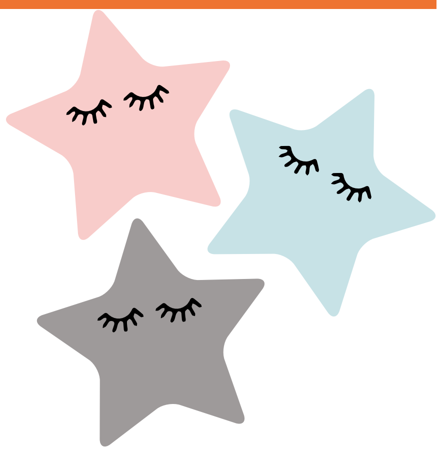

### What will happen on the weekend?

*Friday, Saturday and Sunday*

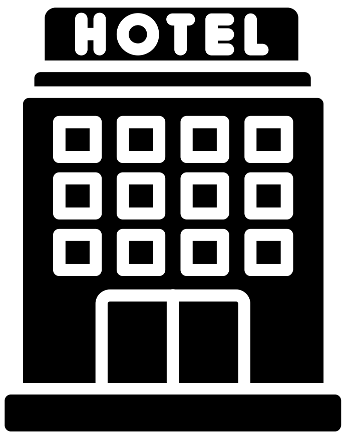


You will travel to Bristol with your families and stay in a hotel.

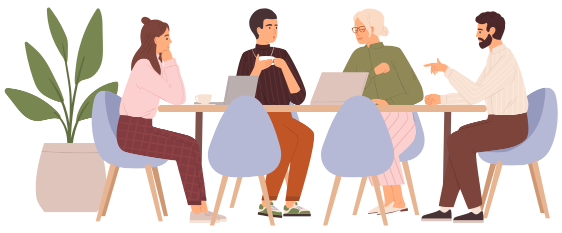

You will come to our sleep space where you will meet some other families who are also being sleep detectives for the weekend.

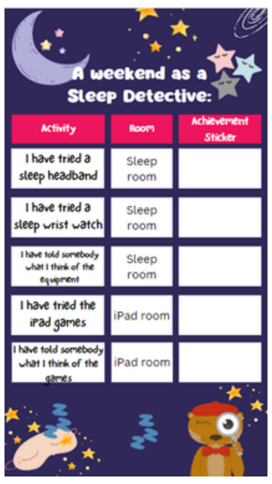

A weekend as a Sleep Detective:

| Activity | Room | Achievement Sticker |
|---|---|---|
| I have tried a sleep headband | Sleep room | |
| I have tried a sleep wrist watch | Sleep room | |
| I have told somebody what I think of the equipment | Sleep room | |
| I have tried the iPad games | iPad room | |
| I have told somebody what I think of the games | iPad room | |

The researchers will ask you to try some equipment and games. You will get stickers for completing these activities.

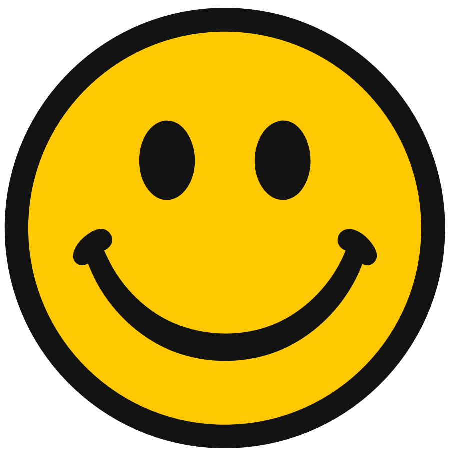

You might try some sleep headbands on, or try a sleep wristwatch. The researcher might ask you to put a headband on and lie back so you can tell if it is comfy.

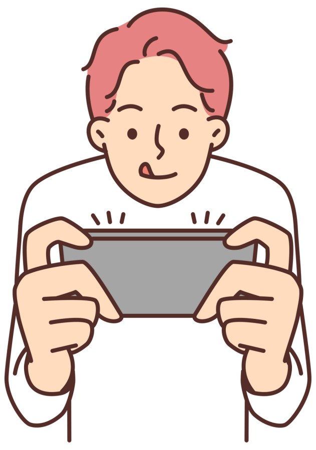

You will also try some games on iPads. The researchers will show you what to do and ask you if you liked playing the games.

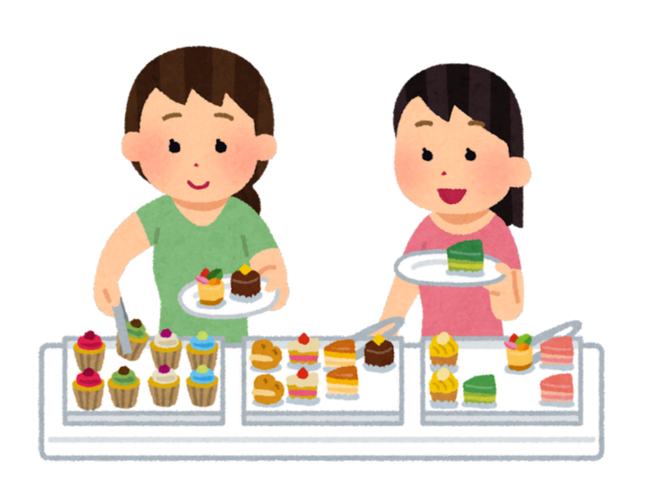

We will have lunch at 1pm - you can help yourselves to what you want to eat from our buffet.

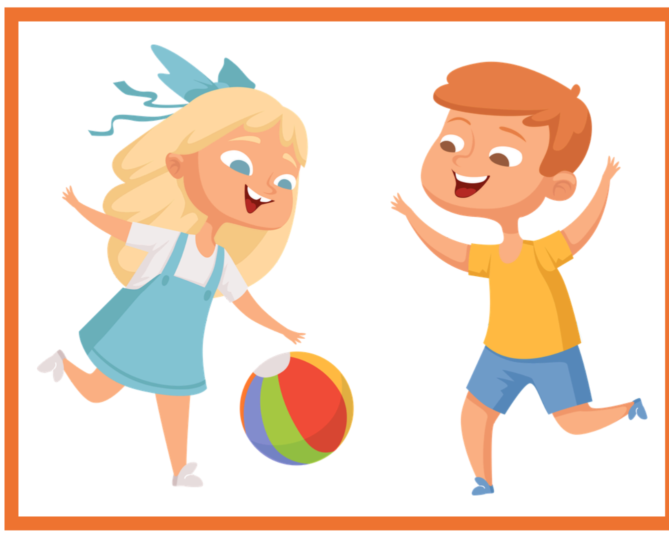

You can also play jigsaws, ball games, lego and colouring. Or you can chill out in our refreshment room or sensory space.

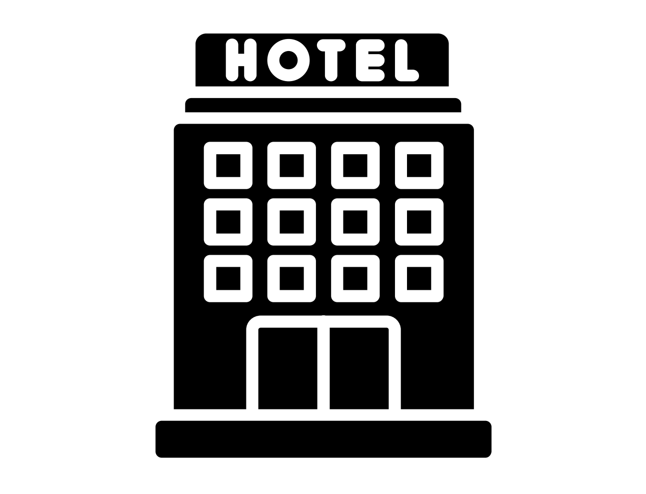


You will stay in your hotel again.

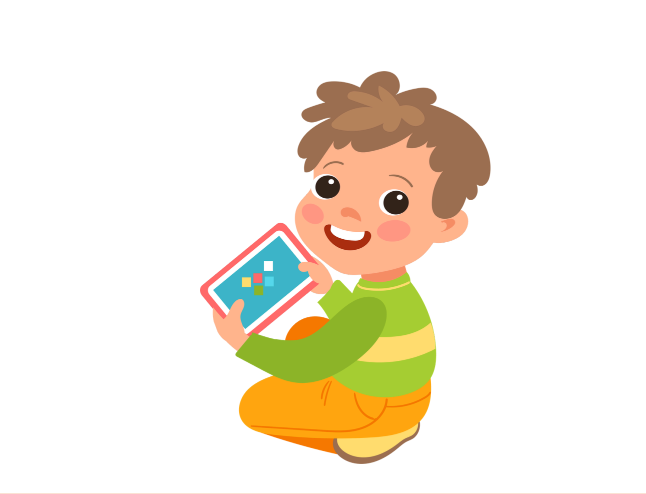

You will come back to the sleep space in the morning to do any activities you didn't have time to do the day before.

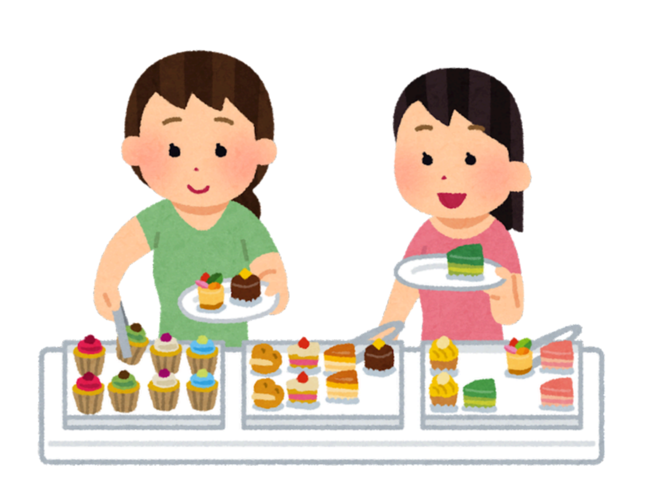

We will have lunch at 12pm - you can help yourselves to what you want to eat from our buffet.

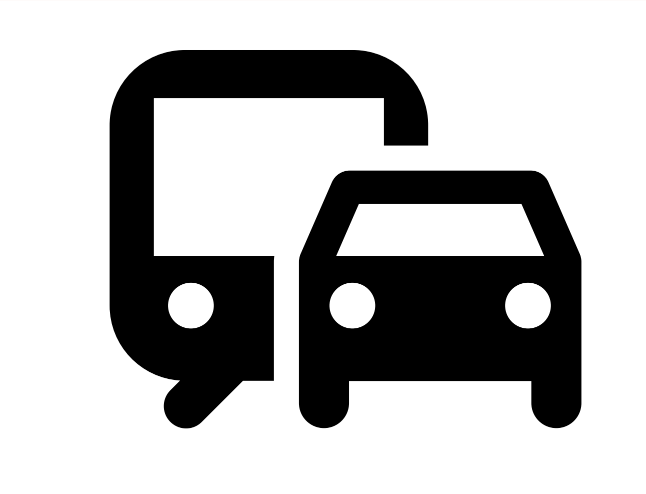

You will go home with your families. Some of you will come in the car, some of you will go on a train.

### Sleep Detectives

Workshop Activity List

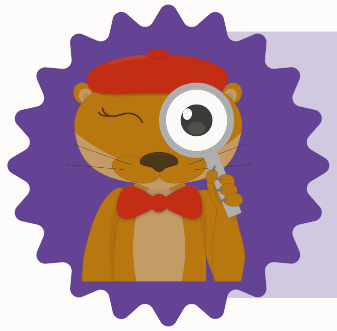

Please take part in any of the activities you like, but remember to help our researchers, by being sleep detectives. Try to complete all the activities on your sticker chart.

| | | | | | | | | | |
|---|---|---|---|---|---|---|---|---|---|
| **Activity** | TRY OUT: Sleep headbands and sleep wristbands | TRY OUT: iPad Games | Design a character: drawing and colouring | Indoor ball play | Quiet sensory rooms: cushions, soft toys, books, calming music | Hands on quiet games: jigsaws, lego | Outdoor play: bean bag throw, coloured chalks, skittles, bubbles | Lunch 1-2pm | Private 1:1 chats |
| **Pictures** | 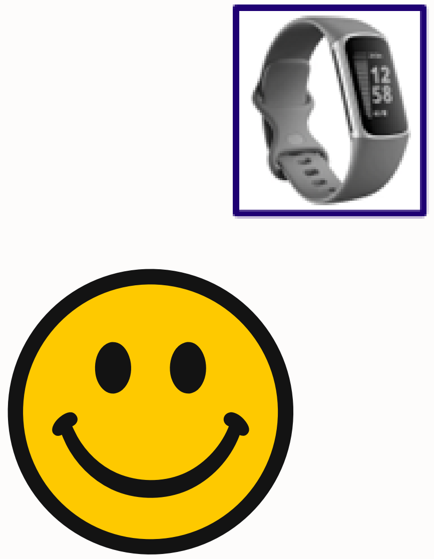 | 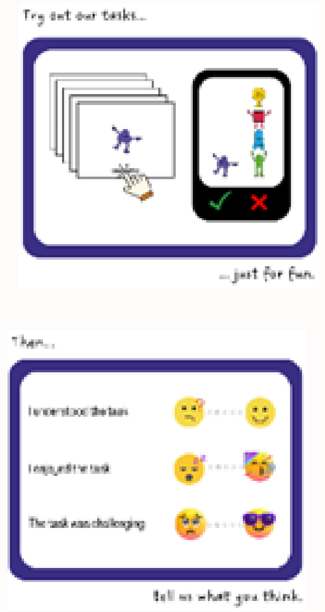 | 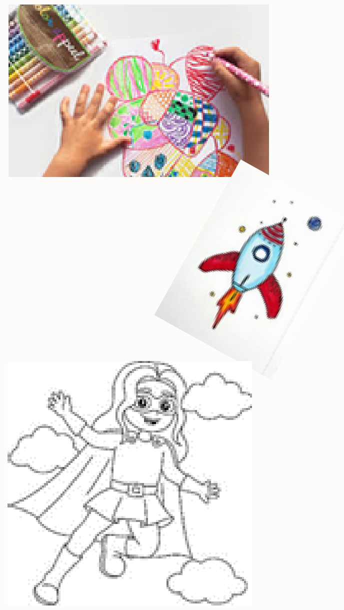 | 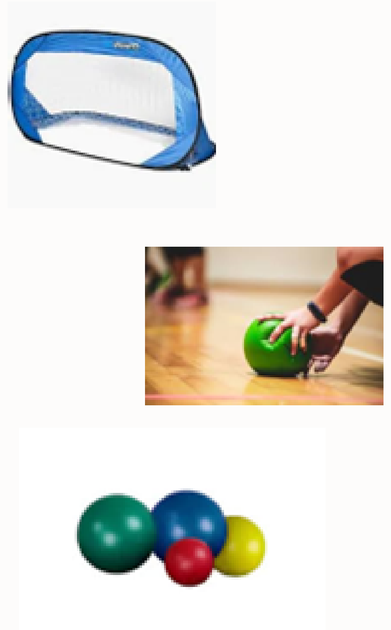 | 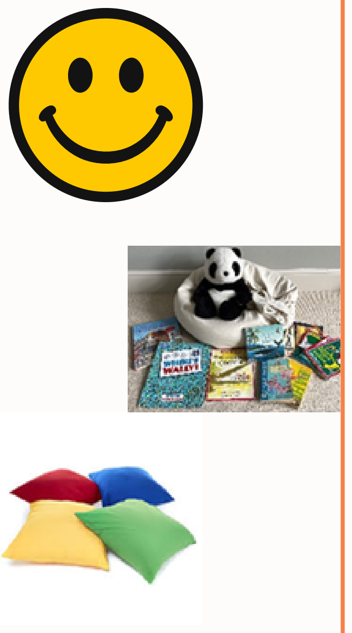 | 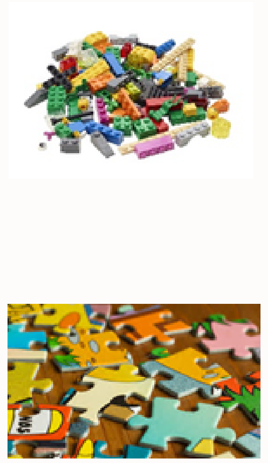 | 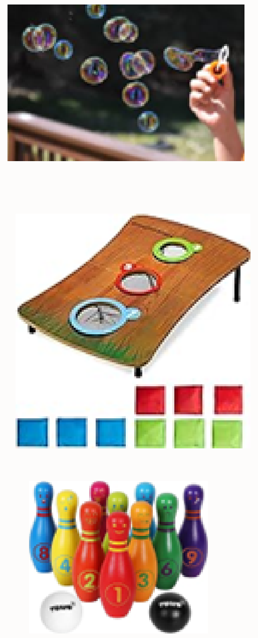 | 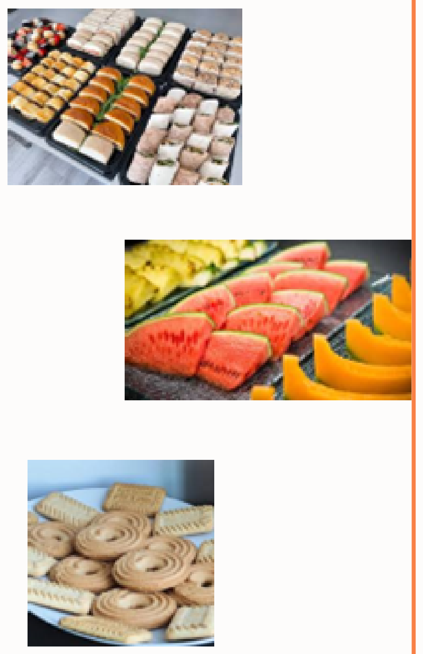 | 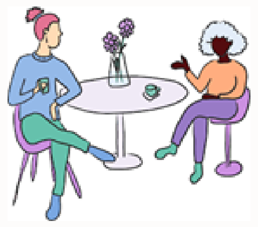 |
| **Room** | Sleep room Lower ground | iPad room Lower ground | Refreshments Area - Lower Ground | Indoor ball room Lower Ground | Sensory room Ground floor | Hands on play room Ground Floor | Outside Yard Lower Ground | Tea Room and Outside Deck Ground Floor | G15 Ground Floor OR LG.23 Lower Ground |

[illegible]

### A weekend as a Sleep Detective:

| Activity | Room | Achievement Sticker |
|---|---|---|
| I have tried a sleep headband | Sleep room | |
| I have tried a sleep wrist watch | Sleep room | |
| I have told somebody what I think of the equipment | Sleep room | |
| I have tried the iPad games | iPad room | |
| I have told somebody what I think of the games | iPad room | |

### Canynge Hall - Bristol

Here is a floor plan of the building where our sleep detectives workshop will take place. This is a sensory map so you can see which rooms might be noisy, or have bright lights, and so you can see the quiet spaces to relax.

#### Ground Floor

1. You will enter onto this floor of the building.
2. NAME and NAME will be on reception (see their pictures)
3. Rooms G11 and G12 will be quiet spaces where people can relax
4. Room G12 will be a quiet multisensory space
5. Room G11 will be a quiet space where you can do activities like lego, and jigsaws
6. The tea room and refreshments area will be a quiet place where people can chat amongst themselves
7. Office G15 can be used as a quiet private space if you need to get out of the hustle and bustle.

Shhh! Quiet voices

Chatter

Natural Light

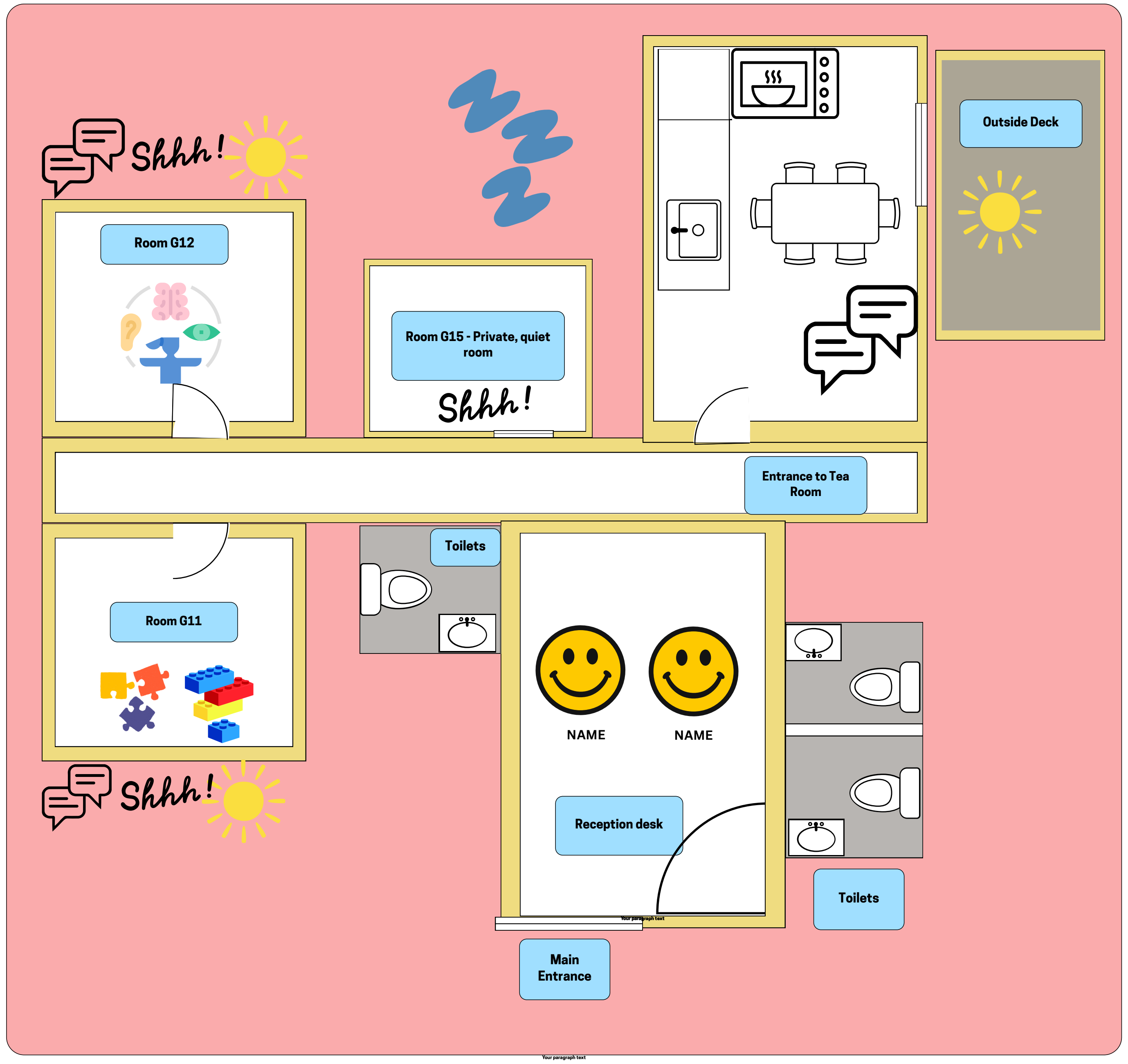


#### Lower Ground Floor

1. You will take stairs down to this level (there is a lift available)
2. Room LG.01 will be used for....
3. Room LG.03 will be our busy activity space - there will be some ipads to play on, and some arts and crafts activities
4. The outside yard will be a safe space for a run around and some ball games - it may be noiser here
5. Room LG08 A and B will be our detective rooms where you can try out our sleep equipment and games

Shhh! Quiet voices

Chatter

Noisy

Natural Light

Bright overhead lights

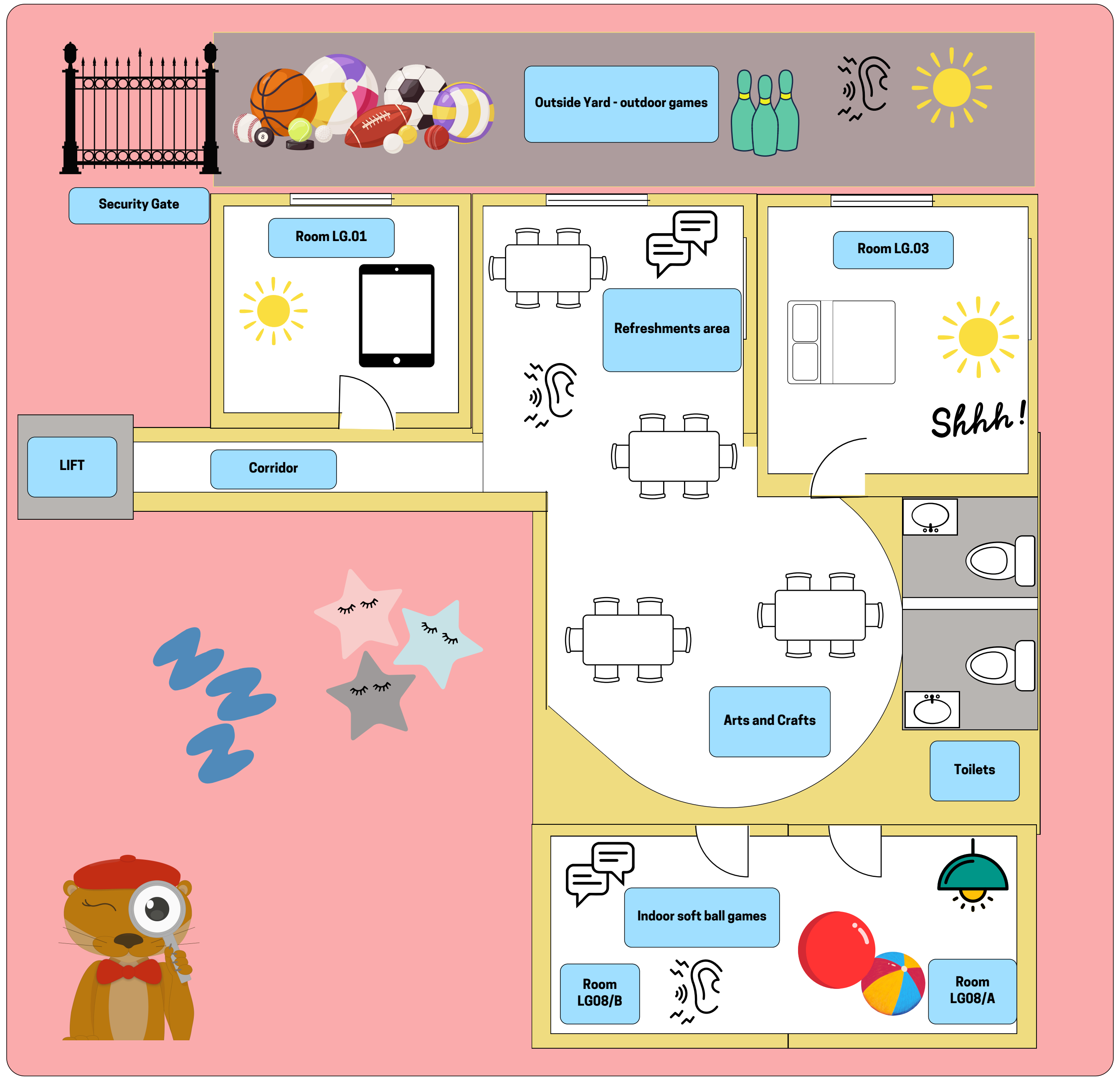


University of BRISTOL

SLEEP DETECTIVES

CARDIFF UNIVERSITY

### Who might I meet?

Here are some of the team, who you might meet on the day!

#### Researchers

NAME | NAME | NAME | NAME | NAME

NAME | NAME | NAME | NAME | NAME

NAME | NAME | NAME | NAME | NAME

#### Charities

NAME

#### Canynge Hall Reception

NAME
